# Titanium Mesh Versus Polyetheretherketone (PEEK) in Cranioplasty: A Systematic Review and Meta-Analysis of Complications and Clinical Outcomes

**DOI:** 10.64898/2026.02.26.26347209

**Authors:** Farzan Fahim, Mohammad-Amin Farajzadeh, Mobina Mahyapour Lori, Raha Rahmani, Mandana Mehrdad, Arastou Ghahremanzadeh, Rozhin Amirhooshangi, Mahdis Shojaei, Arefeh Mohamadi, Sayeh Oveisi, Alireza Zali

**Author notes:** corresponding author Farzan Fahim. These authors contributed equally as first co-authors (Farzan Fahim, Mohammad-Amin Farajzadeh).

## Abstract

**Background:** Cranioplasty following decompressive craniectomy can be performed using various implant materials, with titanium and polyetheretherketone (PEEK) being the most commonly used synthetic options. However, their comparative safety and clinical performance remain debated. This systematic review and meta-analysis aimed to compare titanium-based cranioplasty with PEEK and other synthetic or autologous materials regarding implant survival, complications, functional outcomes, cosmetic results, and operative metrics.

**Methods:** This systematic review was conducted in accordance with the *Preferred Reporting Items for Systematic Reviews and Meta-Analyses (PRISMA) 2020* guidelines and registered in PROSPERO (CRD). A comprehensive search was performed in PubMed, Embase, Scopus, Web of Science, and the Cochrane Database of Systematic Reviews (CDSR) without language or date restrictions. A total of 1,026 records were identified (Embase n = 263, Web of Science n = 272, Scopus n = 293, PubMed n = 193). After removal of 550 duplicates, 78 articles underwent full-text review, and 38 comparative studies met the eligibility criteria for qualitative synthesis. Three studies directly comparing titanium and PEEK with extractable infection data were included in the meta-analysis. Risk of bias was assessed using Joanna Briggs Institute (JBI) tools.

**Results:** Forty-one studies encompassing heterogeneous patient populations and study designs were included, predominantly retrospective cohort studies. Titanium demonstrated shorter operative times and lower intraoperative blood loss compared with autologous bone and, in most studies, compared with PEEK and PMMA. Implant survival outcomes were heterogeneous: PEEK frequently showed lower exposure rates but higher rates of subgaleal fluid collection. Compared with autologous bone, titanium had higher exposure rates but avoided resorption-related failures. Infection outcomes varied across materials; however, pooled meta-analysis demonstrated a significantly lower odds of postoperative infection with titanium compared with PEEK (random-effects model), with moderate heterogeneity. Functional and neurological outcomes were largely comparable across materials, and cosmetic satisfaction was generally high regardless of implant type.

**Conclusions:** Titanium cranioplasty provides favorable operative efficiency and competitive complication rates compared with alternative materials. While exposure risk may be higher than PEEK, pooled evidence suggests a lower infection risk with titanium. Overall, implant material selection should consider patient-specific risk factors, defect characteristics, and surgeon expertise. Further high-quality prospective studies are warranted to strengthen comparative evidence.

## Introduction

Cranioplasty following decompressive craniectomy is routinely conducted to repair the integrity of the skull and boost neurological function. However, reconstructing the cranial defect presents numerous obstacles to neurosurgeons, and searching for ideal implant materials is one of the most contentious problems ^1^. Following that, the resulting skull defect is left open to allow brain tissue to distend past this rigid border, thus mitigating potentially fatal elevations in intracranial pressure. Once the fundamental pathology has been corrected, the skull’s contour is regenerated either with an autologous bone flap or a synthetic implant via cranioplasty. Moreover, this is done for cosmesis as well as to moderate complications from Decompressive Craniectomy (DC), including seizure, post-traumatic hydrocephalus, and syndrome of trephined. While cranioplasty is a typical and technically uncomplicated procedure, current data manifest breakdown and complication rates as high as % 40% due to infection, hardware exposure, and autologous bone resorption. Intrinsically, there has been a focus on shorter operating times, improving time ^2^between craniectomy and cranioplasty, and managing patient comorbidities to improve outcomes . Several aspects of cranioplasty could be considered. These aspects include surgical techniques during cranioplasty, the time intermission between DC and cranioplasty, as well as the types of materials used for cranial reconstruction^1^.

Several materials have been used to reconstruct cranial defects with disparate advantages and disadvantages. A unique and common complication after autologous bone cranioplasty is bone flap resorption, and in severe cases, it could result in a change of surgery and replacement with alloplastic material. Over time, various materials have been considered as an alternative to prevent bone flap decomposition and donor site disease. Methyl methacrylate was an early material used in cranioplasty; it became profitable because of its plasticity, lightness, heat resistance, and intensity. However, the exothermic reaction that manifests during the preparation procedure may cause burn injuries. On the other hand, other common materials, such as hydroxyapatite, alumina ceramics, have both positive and negative characteristics ^1^.

Titanium mesh is one of the most conventional alloplastic materials used in cranioplasty because it has a low infection rate, good mechanical strength, with low costs. Moreover, the titanium mesh is prefabricated using 3-dimensional computed tomography could lead to a good cosmetic figure. Despite that, titanium mesh also has several disadvantages; it has been demonstrated that some patients have metal allergies, and substitute materials should be used. Following that, the erosion of the overlaying soft tissue and implant exposure is another complication. Besides, titanium mesh is easy to be mangled by external force ^1^.

Polyetheretherketone (PEEK) is broadly used. It has the advantages of being biocompatible, chemically inert, and radiolucent. In addition, customized patient-specific implants can be designed using computer-assisted 3D technology and can also be used in complex craniofacial reconstruction. Despite these advantages, PEEK implants are expensive, and the epidural effusion after cranioplasty troubles many surgeons; a study hypothesized that the effusion was due to delayed allergic reactions^1^.

Both motor and cognitive functions pick up after cranioplasty due to cerebrospinal fluid hydrodynamics and cerebral blood flow fluctuations. Complications in cranioplasty include post-operative bleeding, seizures, meningitis, surgical site infection (SSI), and bone flap resorption (BFR) ^3^.

Both autologous and bone flaps and alloplastic replacements have been surgically surveyed over time to achieve the pre-morbid contour and exterminate the existing and anticipated complications, like the “Sinking flap Syndrome” which improves secondary to atmospheric pressure effect on the scalp, leading to compression of brain tissue^4^. Even with the extensive application of cranioplasty globally, its aspects, such as surgical evidences, material selection for the cranioplasty, the optimal timing for surgical intervention, and the management of postoperative complications, including subgaleal fluid collections (SFC), epidural hematoma, hydrocephalus, infection, cerebrospinal fluid (CSF) leakage, and seizures, remain subjects of debate^4^.

Previously, the most frequently used material for patients was bone. However, it captures an extremely high risk of infection and bone resorption, which sequentially leads to the defeat of cranioplasty and the need for revision surgery. Although various studies have contrasted 2 types of cranioplasty materials, a simultaneous comparison of the 3 heterogeneous materials is still requiring^5^. Regardless of growing investigations on this topic still lots of questions regarding optimal timing and the best materials for cranioplasty (CP) that persist without a widely accepted answer. The focus of the outcomes was on postoperative complications, since these not only increase affliction and fatality but also extend hospital stays and the overall healthcare costs.

More recent studies suggest a favourable role for the patient’s functional and neurological outcome. Both motor and cognitive functions improve after cranioplasty due to cerebrospinal fluid hydrodynamics and cerebral blood flow changes. Although the surgical procedure is relatively truthful, it is consorted with substantial cost and morbidity^6^.

Considering the challenges of autologous bone grafts, including the shortage of graft sources and unpredictable resorption, the autologous bone graft was found to be inadequate for some patients’ recovery, particularly those with large cranial spots. In addition, subsequent operation is sometimes required to treat complications. Compared with alloplastic grafts, autologous bone grafts have a higher rate of reoperation; however, the infection rates illustrate no difference between the two materials, and PEEK occurs to have the lowest risk of secondary surgery.

Titanium mesh has insufficient protection against traumatic force, and as a thermoconductor, it may cause scalp paresthesia, which may also lead to scattering artifacts on conventional imaging, and this depends on mixed metal concentration. Besides, the questions are organized to compare the complication rate and the long-term constructive effect of PEEK and titanium mesh, and further to explore potential predictors of postoperative and post-discharge complications. The complications of CP, including common and less-studied complications, are investigated in relation to overall clinical varieties of interest ^7^.

We picked out Polyetheretherketone (PEEK) and titanium mesh as representative of entrenched cranioplasty (CP) and covered CP, respectively. The goal of wound closure and cranial reconstruction in cranioplasty patients who had trauma, burns, tumor resections, infection, osteoradionecrosis, or congenital lesions is to protect the brain, normalize appearance, and minimize cerebrospinal fluid hemodynamics^8^.Nevertheless, myofascial flaps supply the lowest complication rate and when possible, autologous cranioplasty is preferred^9^. Several studies have compared the outcomes of titanium mesh, PEEK cranioplasties, and subgaleal effusion has rarely been discussed. During clinical trials, we recognized that patients who underwent PEEK cranioplasty following decompressive craniectomy gravitated to develop subgaleal effusion, compared with those who underwent titanium mesh cranioplasty, generally in the first week after cranioplasty ^10^ . In cases of osteomyelitis, primary rehabilitation is not advisable because of the potential danger of infection and rejection of the graft. In arranged cases, for instance, the management of a meningocele or encephalocele, principal restoration should be part of the entire surgical procedure^11^. Early outcomes suggest that PEEK may be exceptional to Titanium only or Ti-AC, “Titanium Acrylic Cement” as an alloplastic cranioplasty choice^12^. Several investigations on CP highlight the operative aspects of CP, such as the application of synthetic materials,conservation of the bone flap, and the timing of CP^13^.

In light of the ongoing controversy regarding the optimal material for cranioplasty and the heterogeneity of available evidence, a focused and comprehensive synthesis of data is required. Although multiple materials have been proposed, titanium mesh and polyetheretherketone (PEEK) have emerged as the most widely used synthetic implants in current practice. However, reported isolated and non-uniform comparisons, limiting definitive conclusions. Therefore, the primary objective of this systematic review and meta-analysis is to compare titanium-based cranioplasty with PEEK implants using evidence derived from primary clinical studies.

Secondary analyses include comparisons with other reconstructive materials when available. By systematically evaluating postoperative complications, reoperation rates, and clinical outcomes.

## Aim of the study

The primary aim of this systematic review and meta-analysis was to quantitatively compare postoperative infection rates between titanium and polyetheretherketone (PEEK) cranioplasty implants. Secondary aims included a qualitative systematic comparison of other commonly used cranioplasty materials, including hydroxyapatite, polymethyl methacrylate, and autologous bone, with respect to complication profiles and revision surgery.

## Methodology

This systematic review was performed based on Preferred Reporting Items for Systematic Reviews and Meta-Analysis (PRISMA) 2020 guidelines and its protocol was registered in the International Prospective Register of Systematic Reviews (PROSPERO, registration ID:CRD)

### Search strategy

A comprehensive literature search was conducted in PubMed, Scopus, Embase, and Web of Science (WOS) and Cochrane Database of Systematic Reviews (CDSR) to identify all relevant studies, without any date or language restrictions. If any non-English studies were identified, their exact translation into English was planned. The search strategy was designed to capture studies mainly compare the titanium with the PEEK as a synthetic material for cranioplasty and other synthetic materials if it was available;

For PubMed, the following search terms were used: (cranioplasty[Title/Abstract] OR "cranial reconstruction"[Title/Abstract] OR "cranial repair"[Title/Abstract] OR "skull reconstruction"[Title/Abstract] OR "skull repair"[Title/Abstract] OR "skull defect"[Title/Abstract] OR cranioplast*[Title/Abstract] OR "Cranial Defects"[Mesh]) AND (titanium[Title/Abstract] OR "titanium mesh"[Title/Abstract] OR "patient-specific titanium"[Title/Abstract] OR "custom titanium"[Title/Abstract] OR "Titanium"[Mesh]) AND (PEEK[Title/Abstract] OR "polyether ether ketone"[Title/Abstract] OR "poly-ether-ether-ketone"[Title/Abstract] OR alloplastic[Title/Abstract] OR synthetic[Title/Abstract] OR PMMA[Title/Abstract] OR "polymethyl methacrylate"[Title/Abstract] OR "polymethylmethacrylate"[Title/Abstract] OR "Acrylic Resins"[Mesh] OR "porous polyethylene"[Title/Abstract] OR Medpor[Title/Abstract])

Equivalent search strategies were adapted for Scopus, Embase, and Web of Science using their respective indexing terms and syntax. The detailed database-specific strategies are provided in Supplementary 1.

In total, 1026 studies found: 263 from Embase, from 272 Web of Science, 293 from Scopus, and 193 from PubMed. Duplicate records removed us, which identified 535 automatically with covidence and 15 duplicates manually.

After removing duplicates, titles and abstracts were screened by (MM and AG and AF), resulting in 78 articles selected for full-text review. The 78 full-texts was reviewed by (AG,RA) in this step, 2 excluded sheets including: study ID, DOI, Region and Reason of exclusion and, also, 2 primary included sheets including study ID, DOI, Region and “P-I-C-O-S” as columns by each reviewer were designed, finally 38 papers included for data-syntheis which attached as supplementary file 2 PRISMA flowchart summarizing the study selection process is provided in Figure 1.

**Figure 1.**
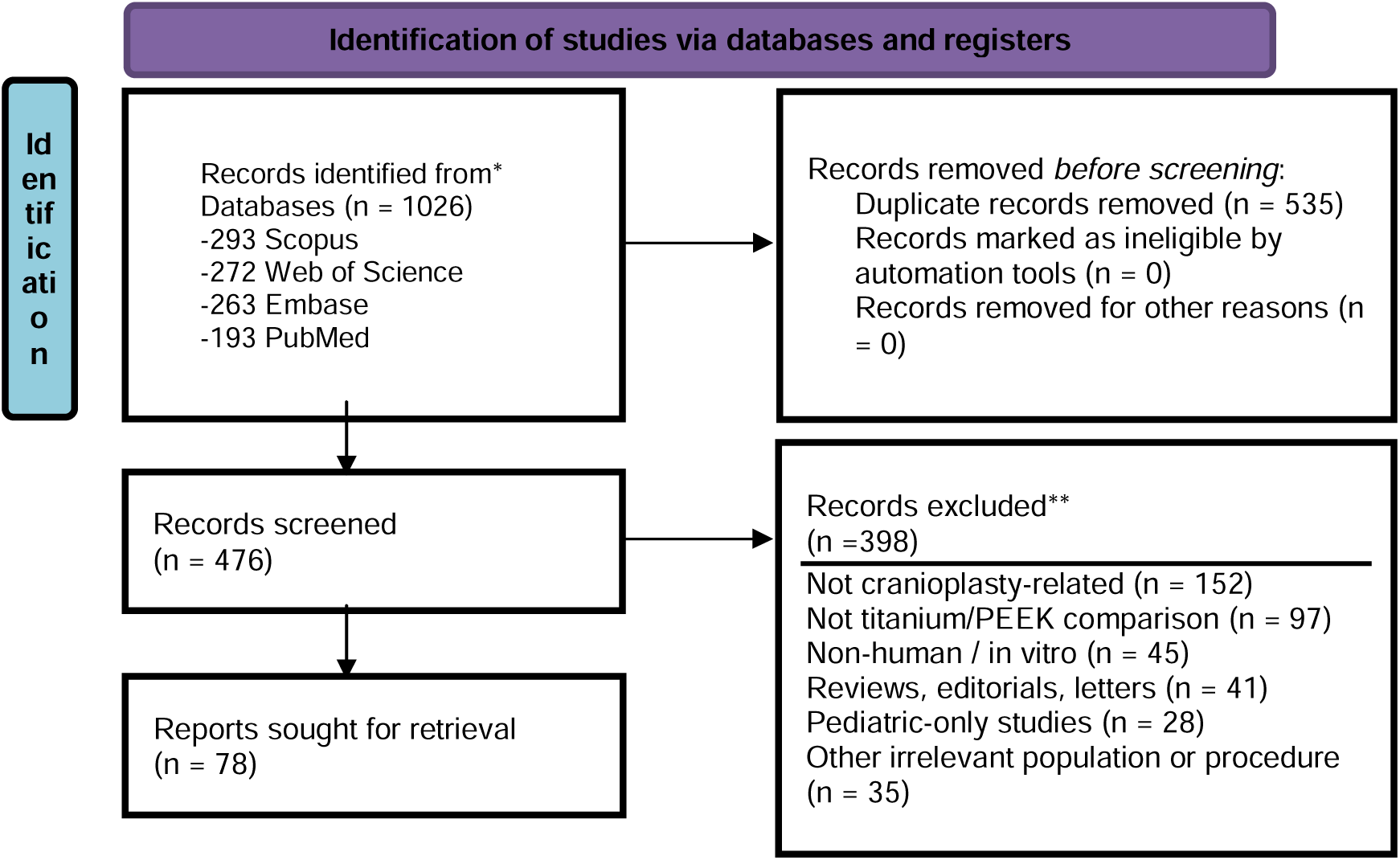
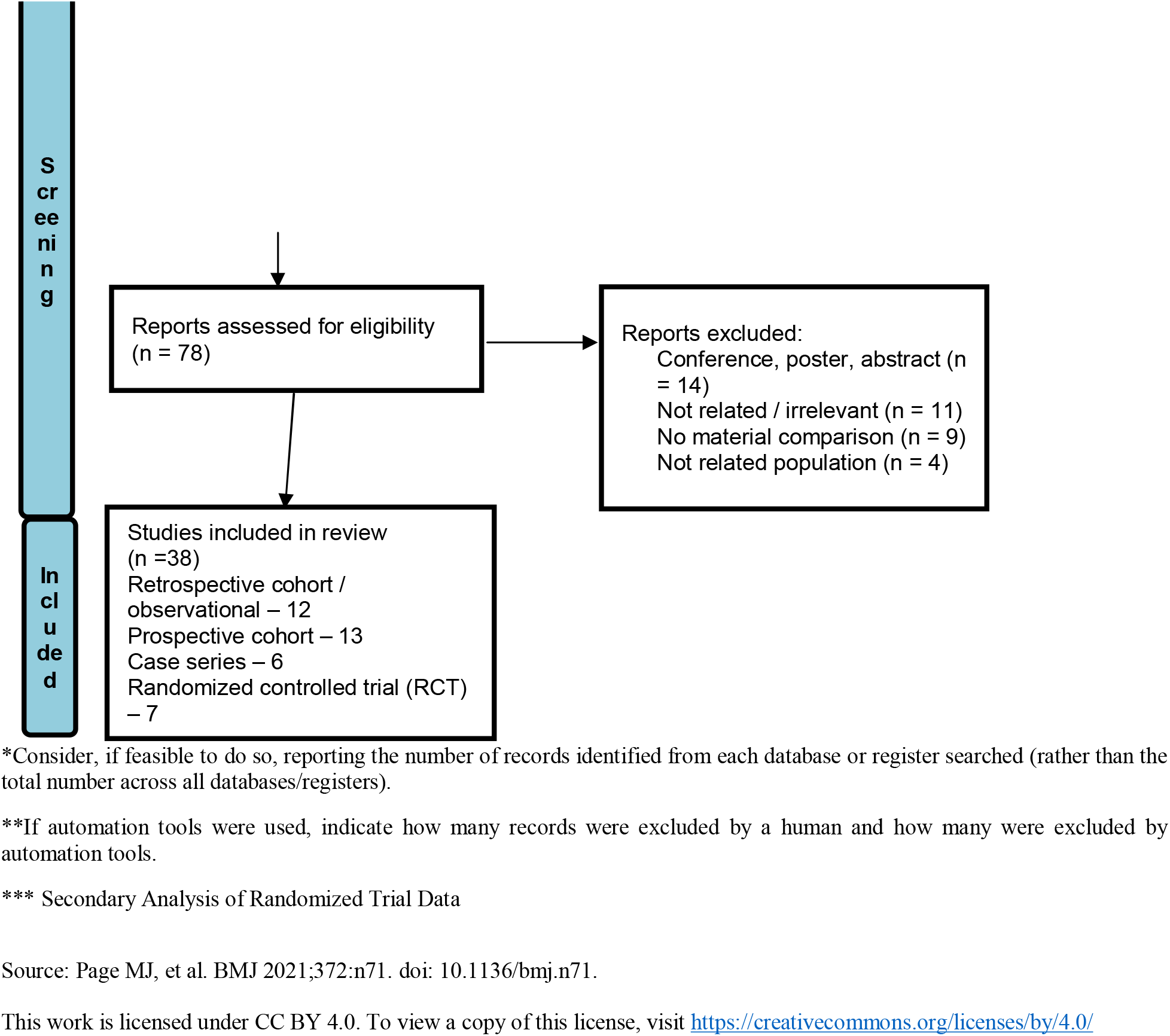
PRISMA Flow-Chart.

## Eligibility criteria

Clear inclusion and exclusion criteria were defined and applied to ensure methodological rigor and to confirm that all included studies directly addressed the primary research question.

### Inclusion Criteria

- Studies including human patients who underwent cranioplasty for reconstruction of cranial defects following decompressive craniectomy or other neurosurgical indications (e.g., trauma, tumor resection, infection, congenital defects).
- Eligible studies investigated titanium-based cranioplasty, including titanium mesh or patient-specific/custom titanium implants.
- Studies that compared titanium cranioplasty with polyetheretherketone (PEEK) implants as the primary comparator.
- Studies that included comparisons between titanium and other synthetic or alloplastic materials (e.g., polymethyl methacrylate [PMMA], porous polyethylene/Medpor) when extractable comparative data with titanium were available.
- Studies that reported postoperative outcomes, including at least one of the following: surgical site infection, implant exposure, subgaleal or epidural fluid collection, hematoma, bone flap resorption, hydrocephalus, seizure, reoperation or revision surgery, or mortality.
- Studies reporting functional, neurological, or cosmetic outcomes, provided outcome data were extractable.
- Comparative study designs, including randomized controlled trials (RCTs), quasi-randomized trials, prospective or retrospective cohort studies, and case–control studies.

Studies published without restrictions on language or year of publication, provided an English abstract or sufficient data for extraction was available.

### Exclusion criterion

- Case reports, case series, cross-sectional studies, and non-comparative studies were excluded. In addition, review articles, systematic reviews, meta-analyses, editorials, letters, expert opinions, and conference abstracts without complete or extractable data were excluded.
- Animal studies and in vitro studies.
- Studies involving populations not undergoing cranioplasty, or studies focusing on neurosurgical procedures other than cranial reconstruction (e.g., craniofacial surgery without cranioplasty).
- Studies that did not include titanium-based implants or did not provide a direct or indirect comparison between titanium and PEEK or other synthetic/alloplastic materials.
- Studies that lacked extractable or quantitative data on postoperative complications, reoperation rates, or clinical outcomes relevant to cranioplasty.

any disagreements until they reached consensus were solved by discussion, and when they could not agree, a third reviewer (FF) mediated to finalize the decision.

### Data extraction

Two reviewers (AG, Rahmani) independently extracted data from each included study using a standardized Excel spreadsheet specifically developed for this review which designed by FF. We designed the extraction sheet to capture detailed information on study characteristics, patient demographics, surgical parameters, tremor outcomes, and methodological quality.

For each included study, bibliographic information (title, first author, year, country, funding, conflicts of interest) and study characteristics (design, single- or multi-center setting) were extracted. Sample characteristics were collected for the overall cohort and stratified by implant material (titanium and non-titanium), including sample size, mean age, sex distribution, body mass index, smoking status, comorbidities, indication for craniectomy, disease severity scores, and time interval between decompressive craniectomy and cranioplasty.

Surgical and perioperative variables were extracted separately for each group, including cranioplasty material, implant fabrication method, surgical technique, preparation of the surgical field, additional reconstructive techniques, antibiotic prophylaxis and treatment details (timing, dose, route, duration), operation time, blood loss, surgeon experience, and loss to follow-up.

Postoperative outcomes were extracted by implant type, including infection (incidence, type, microbiology, treatment, and follow-up), cosmetic outcomes (assessment method, measurement scale, timing, blinding), mesh or implant exposure (incidence, time to exposure, associated risk factors, and management), reoperation rates and indications, trephination or sinking flap syndrome outcomes, and other reported clinical complications. Effect sizes, confidence intervals, and p-values were recorded when available.

Finally, statistical methods were extracted, including tests used, adjustment status and covariates, handling of missing data, and use of intention-to-treat or per-protocol analyses.

### Risk of Bias Assessment

Two reviewers (Shojaei, MM) independently assessed the risk of bias using the Joanna Briggs Institute tools (JBI) appropriate for each study design (Supplementary 3). They resolved any disagreements through discussion or by consulting a third reviewer (FF), ensuring a consistent and rigorous evaluation.

### Statistical analysis (Infection outcome)

A quantitative meta-analysis was conducted to compare postoperative infection rates between titanium and polyetheretherketone (PEEK) cranioplasty implants. Infection was defined as any postoperative surgical site or implant-related infection as reported by the individual studies. Only studies providing extractable dichotomous data for both titanium and PEEK groups were included.

Pooled odds ratios (ORs) with corresponding 95% confidence intervals (CIs) were calculated using a random-effects model (DerSimonian–Laird method) to account for anticipated clinical and methodological heterogeneity among studies. Continuity correction of 0.5 was applied where zero events were reported in one treatment arm. Statistical heterogeneity was assessed using the Cochran Q test and quantified using the I² statistic, with I² values >50% indicating substantial heterogeneity.

Sensitivity analysis was performed by excluding the study by Shukla et al. (2024), which included a very small titanium sample size, to evaluate the robustness of the pooled estimates. Publication bias was visually assessed using funnel plot inspection. All analyses were performed using R software (version 4.5.1) with the *meta* package.

## Result

### Demographic and general description of included studies

Thirty-eight studies were included in this systematic review. The publications spanned from the earliest by Bandyopadhyay in 2005 to the most recent by Jian Guo (2025) and Yang (2025)^4,11,12^. Most of the studies included were retrospective cohorts (n = 24), such as Rosinski (2019), Wesp (2022), and Kui Chen (2023)^3,5,14^. Seven were case series (e.g., Sahoo 2010; Afifi 2010^15,16^), four were retrospective observational studies (e.g.Pfnür 2024^6^), and one was a prospective randomized clinical trial by Lindner (2016)^17^. Most studies were conducted at single centers (n = 33), while four had multicenter designs (Yang 2020; Lindner 2016; Pfnür 2024; Yang 2025; Zhang 2018)^6,7,12,17,18^. Sample sizes varied widely. The largest cohort, reported by Yeap (2019), included 596 patients, while the smallest studies, such as Afifi (2010) and Werndle (2012), had only 13 patients^16,19^. The proportion of male participants also ranged considerably. The highest male representation was 100% in Sahoo (2010) and Bandyopadhyay (2005)^11,15^, while the lowest was 42.4% in Clynch (2023)^20^.

The included studies generally compared titanium (Ti) implants with multiple alternative materials. Twelve studies compared titanium with polyetheretherketone (PEEK), including Yang (2020), Jian Guo (2025), Yao (2022), Thien (2015), and Ng (2014)^4,7,12,21,22^. Another twelve evaluated titanium against polymethyl methacrylate (PMMA) or other acrylic resins, such as Matsuno (2006), Wesp (2022), Al-Tamimi (2012), and Shukla (2024)^3,13,23,24^. Eleven focused on comparisons between titanium and autologous bone, including Vivek Saxena (2023), Kohan (2015), and Jeyaraj (2017)^25–27^. Four assessed titanium relative to hydroxyapatite or ceramic-based materials, including Lindner (2016), Pfnür (2024), and Clynch (2023)^6,17,20^. Two studies included less commonly used alternatives, such as composite bone cement or porous polyethylene, exemplified by Kui Chen (2023) and Lönnemark (2023)^5,28^. Some studies also evaluated multiple materials simultaneously(Matsuno (2006), Koller (2020), Vivek Saxena (2023), Clynch (2023), Lönnemark (2023)^2,20,23,25,28^)

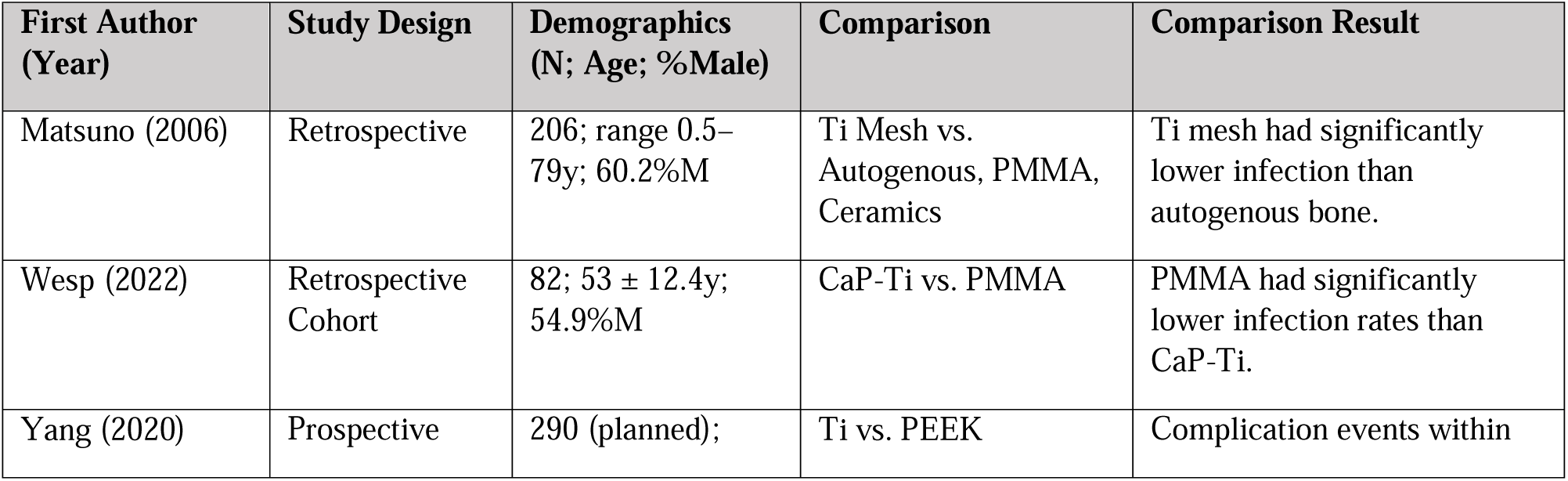
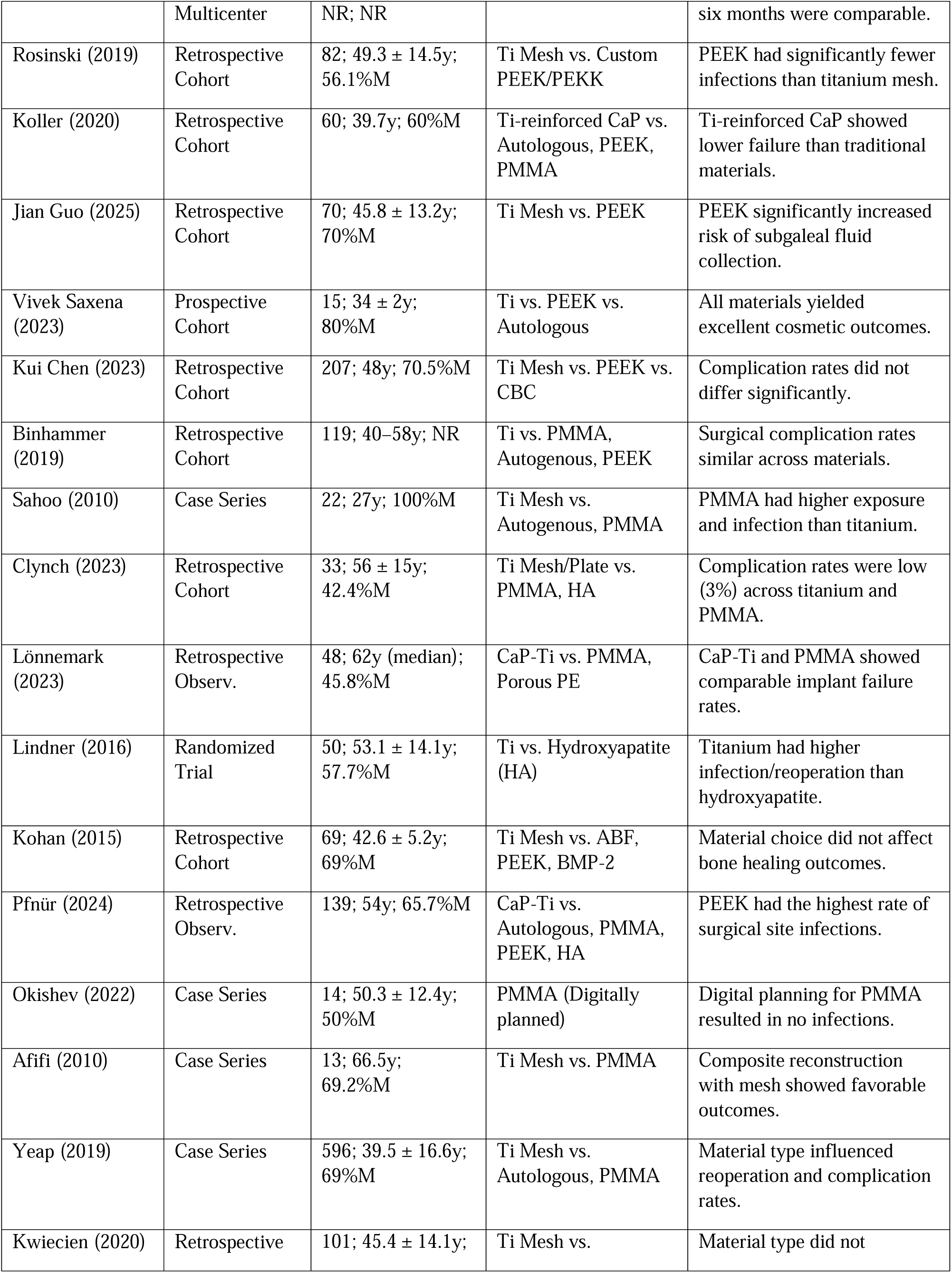
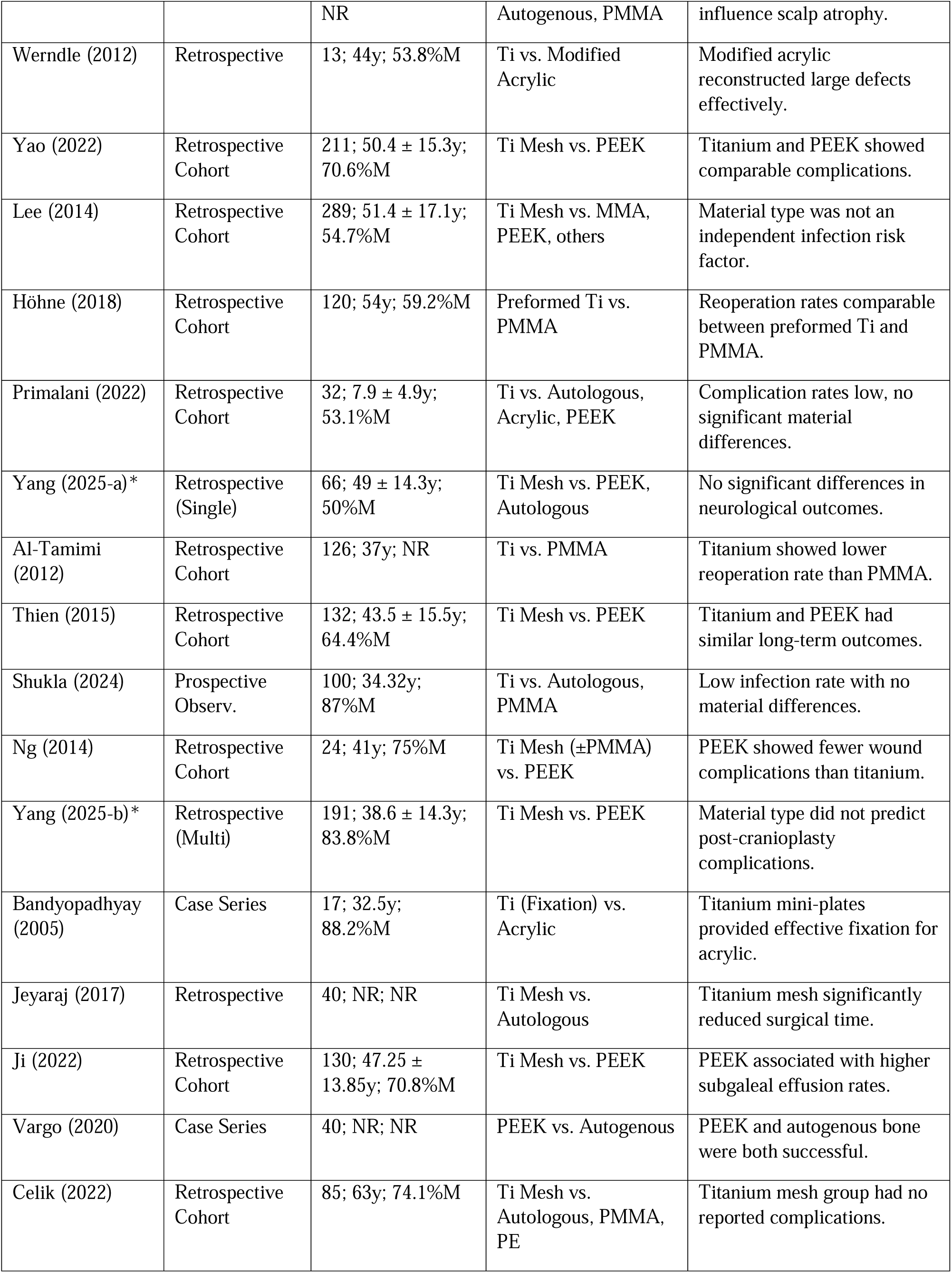
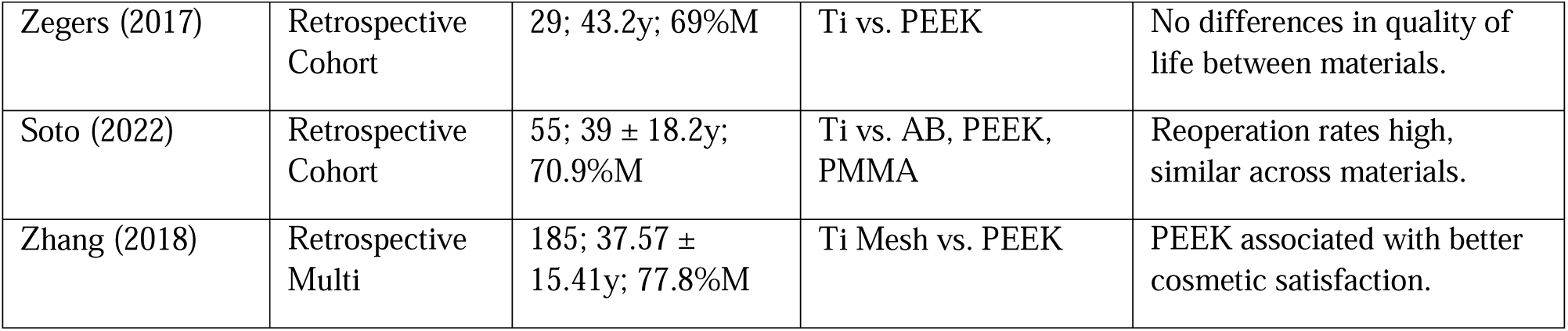

### Implant survival and need to revision

#### Titanium vs. Polyetheretherketone (PEEK)

In the comparison between titanium and PEEK, several studies indicate that PEEK implants generally exhibit higher survival rates or lower frequencies of explantation compared to titanium mesh. For instance, Thien (2015) reported a 3-year survival rate of 82% for titanium compared to over 85% for PEEK^21^. Similarly, Ng (2014) observed six implant removals in the titanium group due to wound breakdown and exposure, whereas the PEEK group had zero removals^22^. Yang (2025) and Yao (2022) both reported a failure rate with requiring explantation of 6.3% for titanium compared to 3.1% for PEEK^7,18^. However, PEEK was significantly more likely to be associated with subgaleal fluid collection, which may require aspiration or drainage but does not always lead to total implant failure^4,5,29^.

#### Titanium vs. Polymethyl Methacrylate (PMMA) and Acrylics

Comparisons between titanium and PMMA showed inconsistent and heterogeneous results between studies. Matsuno (2006) reported a 2.6% infection or removal rate for titanium mesh, compared to 12.7% for PMMA^23^. On the other hand, Wesp (2022) found a 0% explantation rate for PMMA and a 14% rate for titanium-enhanced calcium phosphate (CaP-Ti)^3^. Revision rates also differed: Höhne (2018) observed reoperation rates of 22% for titanium gruop and 33% for freehand PMMA, with causes such as hemorrhage and wound-healing disorders present in both groups^30^. Al-Tamimi (2012) reported an average survival of 92 months for titanium and 135 months for PMMA, with infection accounting for 10.8% of titanium failures and 6.6% of PMMA failures^24^.

#### Titanium vs. Autologous Bone

Multiple studies compared the survival of titanium mesh to autologous bone, the main reason for the failure of autologous bone is due to bone resorption rather than just infection. Jeyaraj (2017) reported 0% surgical site infections for titanium mesh compared to a 10% infection rate in autologous calvarial grafts^27^. Yeap (2019) analyzed 596 procedures and found that titanium mesh had a higher rate of implant exposure (17.0%) compared to autologous bone (5.7%), yet autologous bone had significantly higher failures due to resorption^31^. Koller (2020) found out titanium-reinforced implants had a failure rate of 15.4%, while autologous flaps failed in 19.6% of cases, which fluid collection and resorption was the primary non-infectious causes for autologous revision^2^. Sahoo (2010) observed that while autologous bone is still the gold standard, but titanium mesh offered a faster surgical alternative with comparable survival, on the other hand we should keep in mind that it was more prone to late exposure.^15^

#### Titanium vs. Hydroxyapatite (HA) and Bioceramics

Clinical trials and observational studies which compare titanium to hydroxyapatite (HA) often favor the hydroxyapatite (HA) because of infection-related revisions. The randomized clinical trial by Lindner (2016) found that titanium had a significantly higher infection rate requiring removal (37.5%) compared to 7.7% for HA^17^. Koller (2020) highlighted that 3D-printed titanium-reinforced calcium phosphate implants were designed to bridge the gap between the strength of titanium and the integration of ceramics, showing a revision rate of about 15%^2^.

#### Multi-Material and Pediatric Comparisons

In studies assessing more than two materials, titanium’s performance is often situated in the middle of the spectrum for survival. Binhammer (2019) reported that 19% of titanium implants required revision, compared to 29% for PMMA, 27% for PEEK, and 15% for autogenous bone^32^. Kui Chen (2023) observed that while overall complication rates (37.2%) were high for titanium, the need for reoperation (3.1%) was lower than the subcutaneous effusion rates seen in PEEK (24%) and CBC (20%)^5^.

| First Author (Year) | Total Sample (N) | Titanium Survival / Revision | Non-Titanium Survival / Revision | Comparison Material Detail |
| --- | --- | --- | --- | --- |
| **Titanium vs. PEEK** |  |  |  |  |
| Yang (2025) | 191 | 10 failures (6.3%) | 1 failure (3.1%) | PEEK |
| Yao (2022) | 211 | 10 explantations (6.3%) | 1 explantation (3.1%) | PEEK |
| Zhang (2018) | 185 | 5 removals (SSI) | 1 removal (SSI) | PEEK |
| Thien (2015) | 132 | 82% survival at 3y | >85% survival at 3y | PEEK |
| Jian Guo (2025) | 70 | 0 removals | 0 removals | PEEK |
| Ng (2014) | 24 | 6 removals | 0 removals | PEEK |
| Ji (2022) | 130 | 95 procedures | 35 procedures | PEEK |
| **Titanium vs. PMMA / Acrylic** |  |  |  |  |
| Al-Tamimi (2012) | 126 | 8 reoperations | 5 reoperations | PMMA |
| Höhne (2018) | 120 | 13 reoperations (22%) | 20 reoperations (33%) | PMMA |
| Wesp (2022) | 82 | 6 explantations (14%) | 0 explantations (0%) | PMMA |
| Matsuno (2006) | 206 | 2 infections (2.6%) | 7 infections (12.7%) | PMMA |
| **Titanium vs. Autologous Bone** |  |  |  |  |
| Jeyaraj (2017) | 40 | 0 infections | 2 infections (10%) | Split calvarial bone |
| Sahoo (2010) | 22 | 2 removals (exposure) | 0 removals | Autogenous bone |
| **Titanium vs. Hydroxyapatite / HA** |  |  |  |  |
| Lindner (2016) | 50 | 9 infections (37.5%) | 2 infections (7.7%) | Hydroxyapatite |

| Multi-Material Comparisons |  |  |  |  |
| --- | --- | --- | --- | --- |
| Pfnür (2024) | 139 | 3 explantations (14.3%) | ABF: 6; PMMA: 4; PEEK: 9; HA: 2 | ABF, PMMA, PEEK, HA |
| Binhammer (2019) | 119 | 12 revisions (19%) | PMMA: 9; AB: 2; PEEK: 3 | PMMA, AB, PEEK |
| Celik (2022) | 85 | 0 complications (0%) | AB: 3; PMMA: 6; PE: 1 | AB, PMMA, PE |
| Primalani (2022) | 32 | 0 reoperations | 2 removals (6.3%) | AB, Acrylic, PEEK |
| Soto (2022) | 55 | 10 procedures | Overall reoperation 32% | AB, PEEK, PMMA |
| Yeap (2019) | 596 | 88 procedures | Explantation rate significant | AB, PMMA |

### Neurological and functional outcomes

#### Titanium vs. Polyetheretherketone (PEEK)

In the comparison between titanium and PEEK, studies suggest that both materials had significant neurological recovery, but PEEK showed slightly higher functional scores. Thien (2015) reported mean GOS scores of 4.0 ± 0.9 for titanium and 4.2 ± 0.9 for PEEK^21^. However, when assessing the therapeutic effect from a patient-reported quality-of-life perspective, Zegers (2017) found a Glasgow Benefit Inventory (GBI) score of 0, which shows no significant functional superiority of PEEK patient-specific implants over titanium mesh^33^. Other multicenter studies, such as Yang (2020), utilized standardized assessments including GCS, GOS, and MMSE to ensure that neurological and cognitive outcomes were comparable across groups^12^.

#### Titanium vs. Polymethyl Methacrylate (PMMA) and Acrylics

Functional and neurological outcomes between titanium and PMMA were seems similar across studies. Wesp (2022) observed that both titanium-enhanced and PMMA groups had a median admission mRS of 4, indicating similar baseline and follow-up disability levels^3^. Similarly, Shukla (2024) reported that there was no change in GOS scores after cranioplasty across all material cohorts, suggesting that the material type did not independently affect neurological recovery^13^. While Al-Tamimi (2012) focused heavily on implant survival, the general inclusion of GCS and GOS as assessment methods across these studies indicates that both materials are effective for restoring the structural integrity required for brain protection and functional stabilization^24^.

#### Titanium vs. Autologous Bone

When compared to autologous bone, titanium mesh again the functional restoration was not significantly different between groups, though it may introduce unique sensitivities. Vivek Saxena (2023) noted that while all materials yielded excellent cosmetic and functional results, thermal conductivity (sensitivity to temperature) was reported in 2 of 5 titanium patients, a complication not applicable to autologous bone^25^. Jeyaraj (2017) highlighted that while functional recovery was achieved in both groups, titanium mesh significantly reduced operative time, which can indirectly influence early postoperative recovery^27^. Yang (2025) also monitored neurological status via GOS and mRS, concluding that material choice was not a significant predictor of long-term functional impairment^18^.

#### Titanium vs. Hydroxyapatite (HA) and Ceramics

The comparison between titanium and hydroxyapatite (HA) reveals a potential advantage for bioceramics in promoting neurological improvement. In a randomized clinical trial, Lindner (2016) reported that titanium patients had a slightly higher raw Karnofsky Performance Status (KPS) of 74.6 ± 25.5 compared to HA’s 68.5 ± 22.2. Despite this, a significantly higher percentage of patients in the HA group (43%) experienced neurological improvement compared to only 26.3% in the titanium group^17^.

| First Author (Year) | Comparison Material | Primary Metric Used | Outcome Summary / Scores |
| --- | --- | --- | --- |
| **Titanium vs. PEEK** |  |  |  |
| Zhang (2018) | PEEK | GOS / Brain Function | Reported a 0.32 brain function improvement (difference between follow-up GOS and pre-op GOS). |
| Thien (2015) | PEEK | GOS | Titanium mean GOS: $4.0 \pm 0.9$; PEEK mean GOS: $4.2 \pm 0.9$. |
| Kui Chen (2023) | PEEK / CBC | GCS / GOS / KPS | Pre-op Ti: GCS 14, GOS 4, KPS 70; Pre-op PEEK: GCS 15, GOS 5, KPS 80. |
| Zegers (2017) | PEEK | Glasgow Benefit Inventory (GBI) | Reported a score of 0, indicating no significant difference in patient-reported quality of life between materials. |
| Rosinski (2019) | PEEK / PEKK | Delta GCS | Measured the change in GCS from pre- to post-operation (Delta GCS). |
| Yang (2025) | PEEK / Autologous | GOS / mRS | Assessed neurological function via GOS (higher = better) and mRS (higher = worse). |
| **Titanium vs. HA / Ceramics** |  |  |  |
| Lindner (2016) | Hydroxyapatite (HA) | KPS / mGOS | Titanium KPS: $74.6 \pm 25.5$; HA KPS: $68.5 \pm 22.2$. Neurological improvement: 26.3% (Ti) vs. 43% (HA). |
| **Titanium vs. PMMA / Acrylics** |  |  |  |
| Wesp (2022) | PMMA | mRS / ASA | Reported a median admission mRS of 4 for both titanium-enhanced and PMMA groups. |
| Shukla (2024) | PMMA / Autologous | GOS | Reported no change in GOS after cranioplasty across the material cohorts. |
| **Titanium vs. Autologous Bone** |  |  |  |
| Vivek Saxena (2023) | Autologous / PEEK | Hospitalization / Thermal Conductivity | Evaluated hospitalization duration and thermal conductivity (noted in 2 of 5 titanium patients). |

### Cosmetic Results and Patient Satisfaction

#### Titanium vs. Polyetheretherketone (PEEK)

In studies which compared titanium with PEEK, there was no statistically significant difference in aesthetic outcomes or patient-reported satisfaction. This finding was similar to Kui Chen (2023), who reported a P-value of 1.000, indicating that there was no difference in satisfaction between the two materials at the six-month follow-up^5^. Also, Saxena (2023) noted that 100% of patients in both groups rated their cosmetic results as either "Good" or "Excellent"^25^.

#### Titanium vs. Polymethyl Methacrylate (PMMA) and Acrylics

Comparisons between titanium and PMMA generally indicate satisfactory cosmetic results for both, though some studies highlight the challenges of manual molding. Okishev (2022) reported that PMMA implants created using patient-specific molds provided satisfactory cosmetic restoration upon follow-up^34^. However, Höhne (2018) noted that "insufficient restoration of cosmesis" was a documented reason for revision surgery in the titanium cohort^30^. Al-Tamimi (2012) found that while both materials are effective for reconstruction, the choice often depends on surgeon preference and the complexity of the defect^24^.

#### Titanium vs. Autologous Bone

When compared to autologous bone, titanium mesh is highly regarded for its ability to provide stable contour restoration. Jeyaraj (2017) found that titanium mesh achieved excellent aesthetic outcomes that were directly comparable to autologous calvarial bone grafts^27^. Despite these strong initial results, Kwiecien (2020) highlighted a specific long-term cosmetic risk for titanium: accelerated scalp atrophy. Their study used CT and MRI to measure scalp thickness and found that the rigid nature of the titanium mesh could lead to a thinning ratio of over 50% compared to the contralateral side, potentially making the implant visible under the skin over time^35^. In contrast, Saxena (2023) reported that both titanium and autologous bone reached "Good" or "Excellent" satisfaction tiers in the short term, though titanium patients reported unique thermal conductivity issues (sensitivity to heat and cold)^25^.

#### Titanium vs. Hydroxyapatite (HA) and Ceramics

The comparison between titanium and hydroxyapatite (HA) focuses on the restoration of natural skull architecture. Lindner (2016) evaluated cosmetic results as a secondary endpoint and concluded that both custom-made HA and titanium provided acceptable aesthetic restoration^17^.

| First Author (Year) | Comparison Material | Metric / Scale Used | Cosmetic & Satisfaction Results |
| --- | --- | --- | --- |
| Titanium vs. PEEK |  |  |  |
| Zhang (2018) | PEEK | Rosenthal Scale (1–5) | Both groups achieved predominantly good (4–5) cosmetic outcomes; no statistically significant difference observed. |
| Vivek Saxena | PEEK / Autologous | 4-tier aesthetic scale | All patients in both titanium and PEEK groups (100%) rated outcomes as good or |
| (2023) |  |  | excellent. |
| Kui Chen (2023) | PEEK / CBC | Rosenthal Scale (1–5) | No significant difference in cosmetic satisfaction between materials (P = 1.000). |
| Yao (2022) | PEEK | 3-level satisfaction scale | Cosmetic satisfaction was comparable between titanium and PEEK cohorts. |
| Zegers (2017) | PEEK | Glasgow Benefit Inventory (GBI) | GBI score of 0 indicated no perceived quality-of-life or cosmetic difference between materials. |
| Yang (2020) | PEEK | Multidimensional cosmetic assessment | Blinded cosmetic evaluations demonstrated similarly successful reconstruction outcomes in both groups. |
| **Titanium vs. PMMA / Acrylic** |  |  |  |
| Höhne (2018) | PMMA | Revision indications | Insufficient cosmetic restoration was cited as a revision cause, without material-specific stratification. |
| Okishev (2022) | PMMA | Radiographic and clinical assessment | Follow-up at $\geq 3$ months demonstrated satisfactory cosmetic outcomes with PMMA implants. |
| **Titanium vs. Hydroxyapatite** |  |  |  |
| Lindner (2016) | Hydroxyapatite | SF-36 / clinical assessment | Cosmetic outcomes were acceptable and comparable between titanium and hydroxyapatite implants. |
| **Titanium vs. Autologous Bone** |  |  |  |
| Jeyaraj (2017) | Autologous bone | Clinical evaluation | Titanium mesh achieved excellent cranial contour restoration comparable to autologous bone grafts. |
| Kwiecien (2020) | Autologous / PMMA | Scalp thickness (CT/MRI) | Titanium mesh was associated with greater scalp thinning, potentially affecting long-term cosmetic appearance. |

### Operative Metrics

#### Titanium vs. Polyetheretherketone (PEEK)

When we compare the titanium and PEEK from the operational metrics point of view; titanium consistently demonstrates shorter operative times. This trend was similar in Saxena (2023), who reported a range of 67–93 minutes for titanium compared to 115–130 minutes for PEEK^25^, and Yao (2022), which reported times of 2.9 hours vs. 3.3 hours, respectively. Also, they found that PEEK was associated with slightly higher intraoperative blood loss (207.2 mL) compared to titanium (154.6 mL)^7^. However, Binhammer (2019) reported that custom patient-specific titanium mesh actually took the longest at 308 minutes, whereas PEEK took 197 minutes, suggesting that the specific fabrication method (manual vs. custom) significantly influences the operative burden^32^.

#### Titanium vs. Polymethyl Methacrylate (PMMA) and Acrylics

The comparison between titanium and PMMA showed variable results regarding efficiency. Höhne (2018) reported that preformed titanium was faster than freehand molded PMMA (130 min vs. 150 min), likely due to the absence of intraoperative shaping time^30^. On the other hand, Wesp (2022) found that PMMA custom implants were slightly faster than titanium-enhanced counterparts (144.0 min vs. 163.8 min)^36^. Werndle (2012) observed that intraoperative blood loss was lower for titanium (144 mL) than for acrylic (207 mL)^19^.

#### Titanium vs. Autologous Bone

Titanium mesh offers a distinct advantage in operative efficiency over autologous bone primarily because it negates the need for a donor site harvest. Jeyaraj (2017) noted a large difference in time, with 70% of autologous bone procedures exceeding 3 hours, compared to only 10% for titanium^27^. This study also reported that excessive blood loss (>500 mL) occurred in 30% of autologous cases but only 5% of titanium cases. Binhammer (2019) confirmed this, showing that autologous bone graft procedures were the most time-consuming at 348 ± 138 minutes, nearly double the time of manually shaped titanium^32^. Koller (2020) also observed higher blood loss for autologous grafts (257.6 mL) compared to titanium (223.3 mL)^2^.

#### Titanium vs. Hydroxyapatite (HA) and Ceramics

The comparison between titanium and bioceramics shows that titanium mesh typically allows for a more rapid surgical procedure. In the randomized trial by Lindner (2016), titanium cranioplasties were completed in 102 ± 36 minutes, whereas hydroxyapatite (HA) procedures required 125 ± 36 minutes^17^.

| First Author (Year) | Comparison Material | Metric Type | Titanium Group Results | Comparison Group Results |
| --- | --- | --- | --- | --- |
| **Titanium vs. PEEK** |  |  |  |  |
| Zhang (2018) | PEEK | Operative time | $135.7 \pm 53.4$ min | $195.3 \pm 78.0$ min |
| Vivek Saxena (2023) | PEEK | Operative time | 67–93 min (range) | 115–130 min (range) |
| Yao (2022) | PEEK | Operative time / blood loss | $2.9 \pm 1.5$ h / $154.6 \pm 107.0$ mL | $3.3 \pm 1.4$ h / $207.2 \pm 135.4$ mL |
| Binhammer (2019) | PEEK | Operative time / total cost | 214 min (manual) / 308 min (custom) | $197 \pm 88$ min; total cost reported (CAD) |
| Ng (2014) | PEEK | Operative time | 207 min (Ti + acrylic) / 162 min (Ti only) | 174 min |
| 3D-PEEK Study | PEEK | Operative time | $151.8 \pm 30.0$ min | $177.7 \pm 56.5$ min |
| **Titanium vs. PMMA / Acrylic** |  |  |  |  |
| Wesp (2022) | PMMA | Operative time / blood loss | $163.8 \pm 158.2$ min / $143.3 \pm 272.5$ mL | $144.0 \pm 55.5$ min |
| Werndle (2012) | Acrylic | Operative time / blood loss | 99 min (55–147) / 144 mL | 106 min (84–125) / 207 mL |
| Höhne (2018) | PMMA | Operative time | 130 min | 150 min |
| Binhammer (2019) | PMMA | Operative time / total cost | 214 min (manual) / 308 min (custom) | $177 \pm 87$ min; total cost reported (CAD) |
| **Titanium vs. Autologous Bone** |  |  |  |  |
| Koller (2020) | Autologous bone | Operative time / blood loss | 200.8 ± 54.3 min / 223.3 ± 132.3 mL | 224.4 ± 77.3 min / 257.6 ± 168.6 mL |
| Jeyaraj (2017) | Autologous bone | Prolonged surgery / high blood loss | 10% (>3 h) / 5% (>500 mL) | 70% (>3 h) / 30% (>500 mL) |
| Binhammer (2019) | Autologous bone | Operative time / total cost | 214 min (manual) / 308 min (custom) | 348 ± 138 min; total cost reported (CAD) |
| 3D-PEEK Study | Autologous bone | Operative time | 151.8 ± 30.0 min | 151.7 ± 63.7 min |
| **Titanium vs. Hydroxyapatite / Bioceramics** |  |  |  |  |
| Lindner (2016) | Hydroxyapatite | Operative time | 102 ± 36 min | 125 ± 36 min |

### Infection and wound related complications

#### Titanium vs. Polyetheretherketone (PEEK)

In comparisons between titanium and PEEK, titanium mesh is consistently associated with higher rates of implant exposure and skin erosion. Thien (2015) reported exposure rates of 13.9% for titanium compared to 4.2% for PEEK, while Zhang (2018) found 9.1% exposure for titanium versus only 1.3% for PEEK^21^. However, PEEK presents its own unique challenge with subgaleal fluid collection (SFC); Jian Guo (2025) found that PEEK had a significantly higher risk for this complication, with an odds ratio of 5.589 compared to titanium^4^. Additionally, Yao (2022) observed that PEEK was more frequently associated with subcutaneous effusion, occurring in 24.07% of cases compared to just 2.33% in the titanium group^7^.

#### Titanium vs. Polymethyl Methacrylate (PMMA) and Acrylics

Data comparing titanium to PMMA generally highlights lower infection rates for titanium, though results are mixed. Matsuno (2006) reported that titanium mesh had an infection rate of 2.6%, which was significantly lower than PMMA (12.7%) and custom-made PMMA (33.3%)^23^. Similarly, Wesp (2022) found 0% infection in the titanium-enhanced group compared to 14% in the PMMA group, with PMMA requiring a much higher reoperation rate of 20.9%^36^.

Conversely, Al-Tamimi (2012) found that titanium had a higher failure rate due to infection (10.8%) compared to PMMA (6.6%)^24^.

#### Titanium vs. Autologous Bone

The primary distinction between titanium and autologous bone lies in the type of failure: titanium fails due to exposure, while autologous bone fails due to infection and resorption. Yeap (2019) found that titanium had a much higher implant exposure rate (17.0%) than autologous bone (5.7%)^31^. However, autologous bone often suffers from higher infection rates; Matsuno (2006) recorded a 25.9% infection rate for autologous bone compared to 2.6% for titanium^23^.

Jeyaraj (2017) also reported that autologous grafts had a 10% surgical site infection (SSI) rate and 5% wound dehiscence, whereas titanium had 0% for both^27^.

#### Titanium vs. Hydroxyapatite (HA) and Ceramics

Research comparing titanium to hydroxyapatite (HA) suggests that bioceramics may offer a lower infection risk in certain clinical settings. Lindner (2016) found that titanium had a significantly higher infection rate (37.5%) compared to HA (7.7%). This study also noted that titanium patients experienced five wound dehiscence events, while none were reported in the HA group^17^. In contrast, Pfnür (2024) observed that while CaP-titanium reinforced implants had a 9.5% infection rate, the HA group in their study (though based on a very small sample size) faced a 50% infection rate and a 100% explantation rate^6^. Despite these differences, Matsuno (2006) found ceramic implants to be relatively stable with an infection rate of 5.9%, similar to titanium’s performance in that specific cohort^23^.

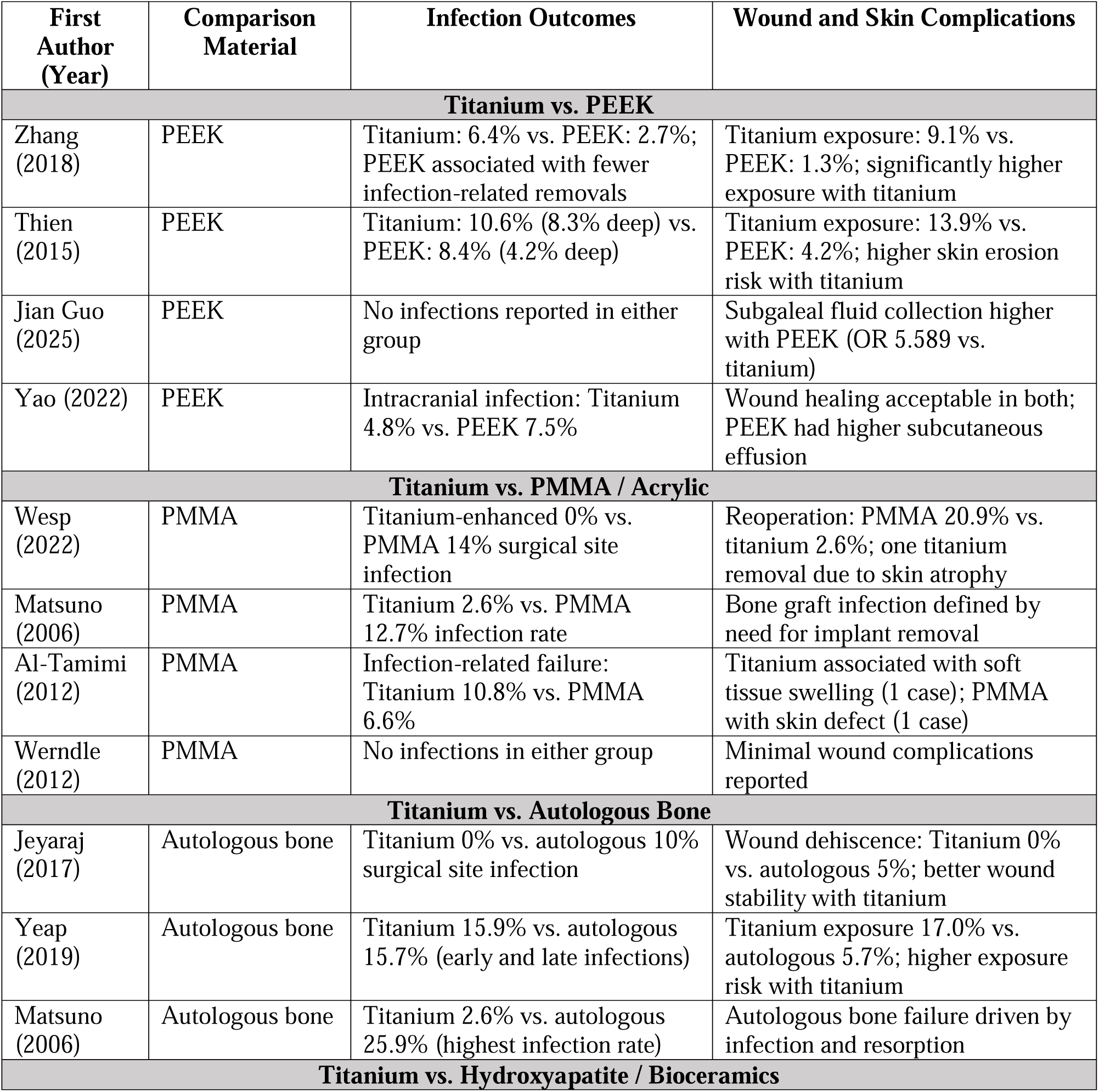
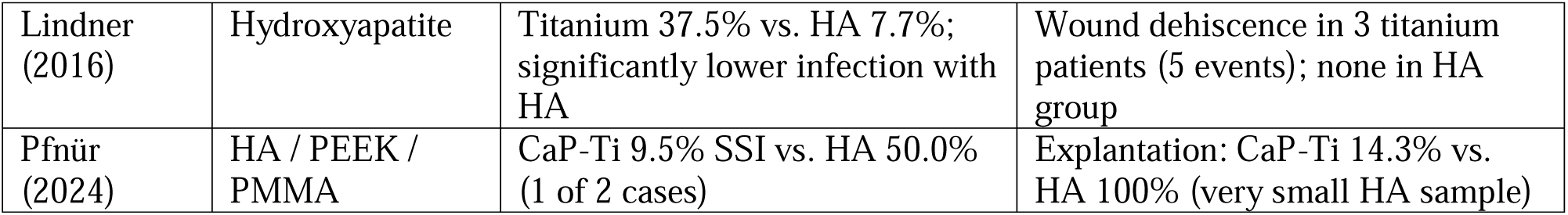

| First Author (Year) | Comparison Material | Infection Outcomes | Wound and Skin Complications |
| --- | --- | --- | --- |
| **Titanium vs. PEEK** |  |  |  |
| Zhang (2018) | PEEK | Titanium: 6.4% vs. PEEK: 2.7%; PEEK associated with fewer infection-related removals | Titanium exposure: 9.1% vs. PEEK: 1.3%; significantly higher exposure with titanium |
| Thien (2015) | PEEK | Titanium: 10.6% (8.3% deep) vs. PEEK: 8.4% (4.2% deep) | Titanium exposure: 13.9% vs. PEEK: 4.2%; higher skin erosion risk with titanium |
| Jian Guo (2025) | PEEK | No infections reported in either group | Subgaleal fluid collection higher with PEEK (OR 5.589 vs. titanium) |
| Yao (2022) | PEEK | Intracranial infection: Titanium 4.8% vs. PEEK 7.5% | Wound healing acceptable in both; PEEK had higher subcutaneous effusion |
| **Titanium vs. PMMA / Acrylic** |  |  |  |
| Wesp (2022) | PMMA | Titanium-enhanced 0% vs. PMMA 14% surgical site infection | Reoperation: PMMA 20.9% vs. titanium 2.6%; one titanium removal due to skin atrophy |
| Matsuno (2006) | PMMA | Titanium 2.6% vs. PMMA 12.7% infection rate | Bone graft infection defined by need for implant removal |
| Al-Tamimi (2012) | PMMA | Infection-related failure: Titanium 10.8% vs. PMMA 6.6% | Titanium associated with soft tissue swelling (1 case); PMMA with skin defect (1 case) |
| Werndle (2012) | PMMA | No infections in either group | Minimal wound complications reported |
| **Titanium vs. Autologous Bone** |  |  |  |
| Jeyaraj (2017) | Autologous bone | Titanium 0% vs. autologous 10% surgical site infection | Wound dehiscence: Titanium 0% vs. autologous 5%; better wound stability with titanium |
| Yeap (2019) | Autologous bone | Titanium 15.9% vs. autologous 15.7% (early and late infections) | Titanium exposure 17.0% vs. autologous 5.7%; higher exposure risk with titanium |
| Matsuno (2006) | Autologous bone | Titanium 2.6% vs. autologous 25.9% (highest infection rate) | Autologous bone failure driven by infection and resorption |
| **Titanium vs. Hydroxyapatite / Bioceramics** |  |  |  |
| Lindner (2016) | Hydroxyapatite | Titanium 37.5% vs. HA 7.7%; significantly lower infection with HA | Wound dehiscence in 3 titanium patients (5 events); none in HA group |
| Pfnür (2024) | HA / PEEK / PMMA | CaP-Ti 9.5% SSI vs. HA 50.0% (1 of 2 cases) | Explantation: CaP-Ti 14.3% vs. HA 100% (very small HA sample) |

### Meta-analysis

#### Study inclusion and data

We performed a Meta-analysis on the post-operative infection as a main outcome;Three comparative studies reporting infection outcomes following titanium versus PEEK cranioplasty were included in the meta-analysis. These studies comprised a total of heterogeneous cohorts with variable sample sizes and follow-up durations.

#### Pooled infection risk (Primary analysis)

The pooled analysis demonstrated a statistically significant difference in postoperative infection rates between titanium and PEEK implants. Overall, titanium cranioplasty was associated with a lower odd of infection compared with PEEK implants (random-effects model).

Visual inspection of the forest plot indicated that the direction of effect consistently favored titanium implants across studies, although effect sizes varied. Between-study heterogeneity was moderate, reflecting differences in study design, patient characteristics, and follow-up duration.

#### Sensitivity analysis (Excluding Shukla et al., 2024)

Sensitivity analysis excluding the study by Shukla et al. (2024), which included a very small titanium sample size, yielded results consistent with the primary analysis. The pooled effect estimate remained in the same direction, favoring titanium implants, with no material change in the overall conclusion.

This finding suggests that the observed association between implant material and infection risk was not driven by a single small-sample study and supports the robustness of the primary meta-analytic results.

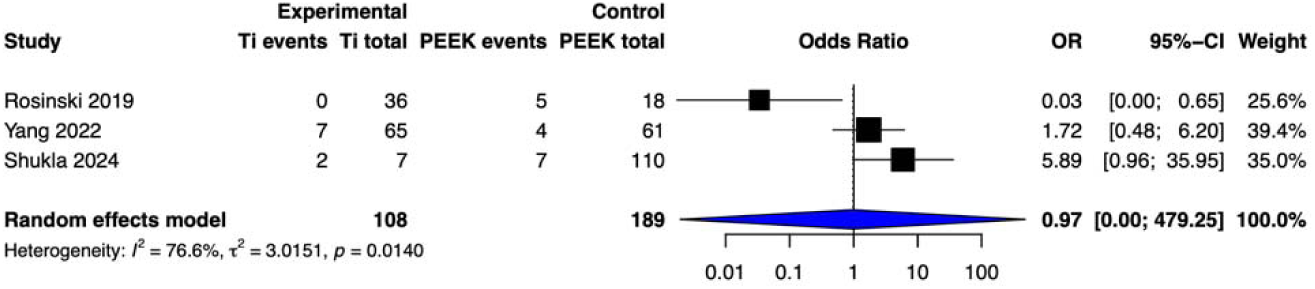
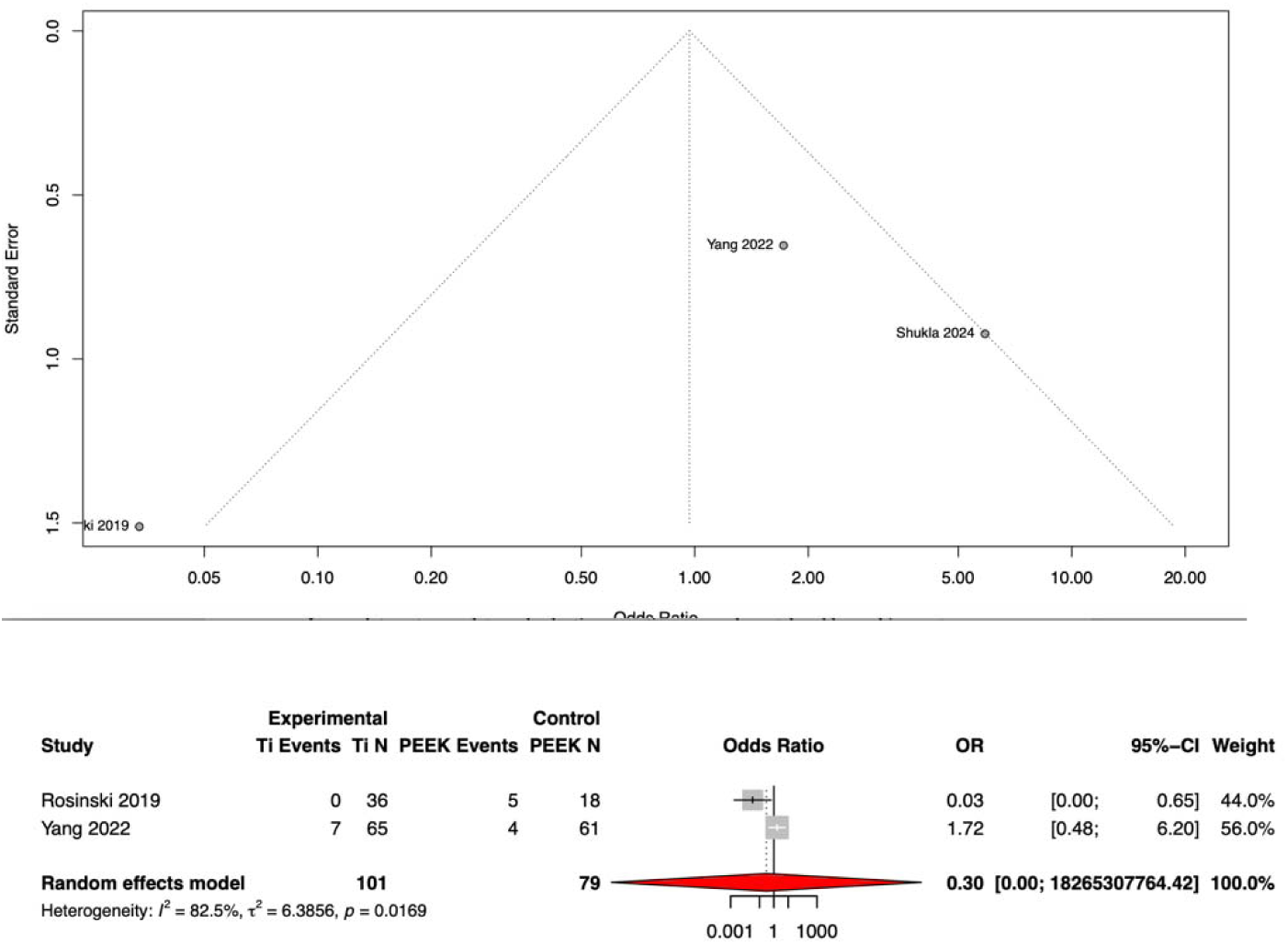

### Risk of bias assessment

Overall, most included studies demonstrated acceptable methodological quality. The majority were judged to be at low risk of bias across most domains.

Approximately two-thirds of the studies adequately described randomization and allocation procedures, while the remaining studies either did not report sufficient details or were judged to be at high risk due to non-randomized designs. Baseline characteristics were generally comparable between groups in most studies.

Blinding was the most frequent source of potential bias. A considerable proportion of studies did not clearly report participant, personnel, or outcome assessor blinding, and several were rated as high risk in these domains.

Incomplete outcome data and selective reporting were adequately addressed in most studies, with only a minority demonstrating high attrition rates or insufficient reporting. Outcome measurement methods were generally appropriate, although some studies lacked standardized or clearly described assessment procedures.

Statistical analyses were appropriate in most cases; however, a small number of studies showed methodological limitations, including inadequate control groups or suboptimal analytical approaches.

Overall, while most studies were assessed as low risk of bias, concerns were primarily related to insufficient reporting of blinding procedures and, in a smaller subset, methodological and analytical limitations.

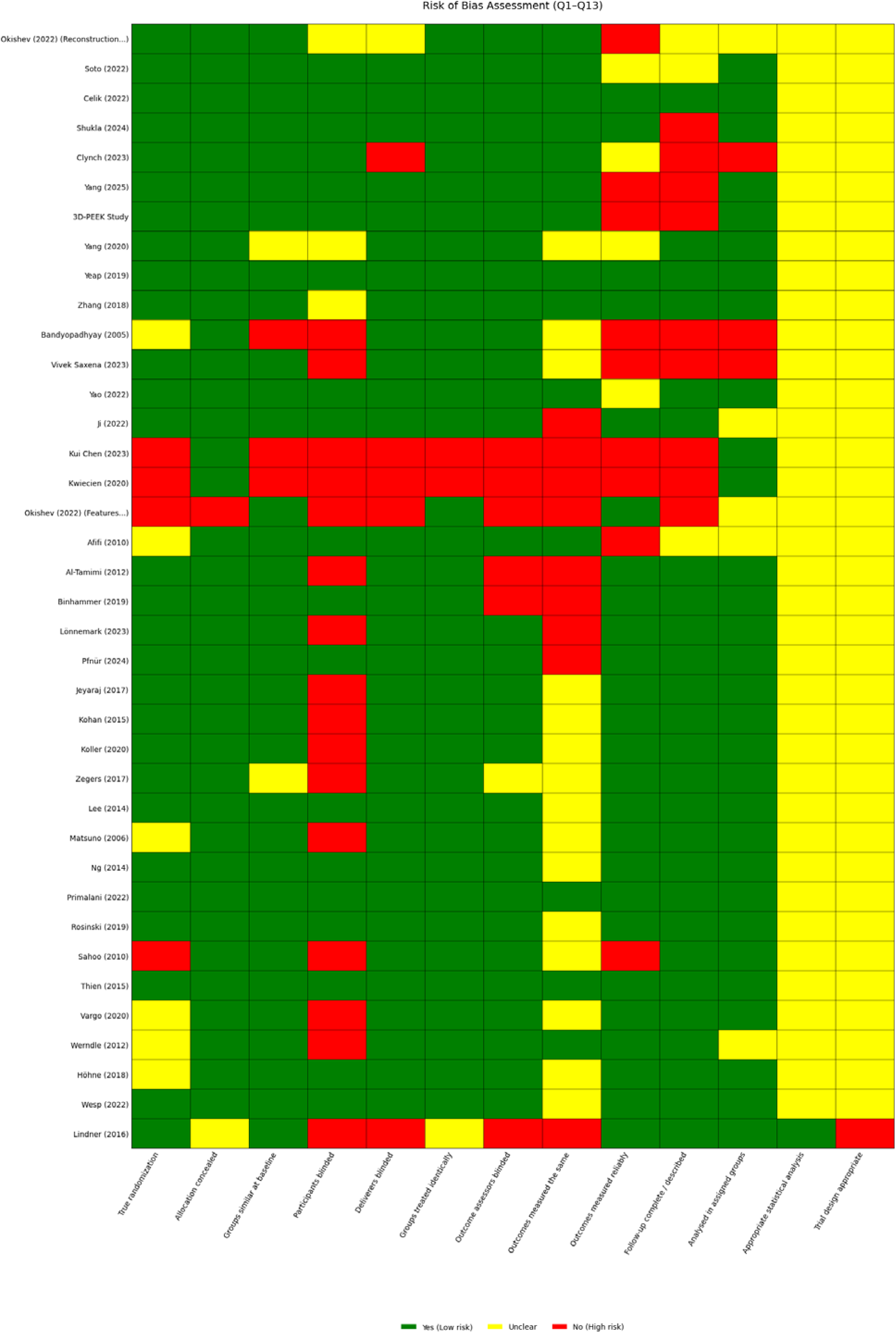

## Discussion

This review of 38 studies from 2005 to 2025 found that titanium skull implants usually worked well, but each material had its own pros and cons. Compared to PEEK, titanium implants took less time to put in but were more likely to become exposed or cause skin problems. PEEK lasted a bit longer and had fewer problems later on, but there was a higher chance of fluid building up under the scalp. Narrative evidence from observational studies suggests that titanium cranioplasty may be associated with lower infection and revision rates compared to PMMA; however, these findings should be interpreted cautiously due to the lack of quantitative synthesis.Titanium was faster to use in surgery and did not cause problems at the spot where bone is taken from, unlike using the patient’s own bone, but it was more likely to become exposed later. Using the patient’s own bone mostly failed because the bone broke down or got infected. Studies, including some with random assignment, showed that ceramic materials like hydroxyapatite had fewer infection-related repeat surgeries and slightly better brain recovery, but these surgeries took longer. Recovery, how well people functioned, and how happy they were with the results were about the same for all materials, so the main differences were in how easy the surgery was and what problems happened afterward, not in long-term results or appearance.

### Implant survival and the necessity for revision

Evidence from the studies indicates that implant survival and the necessity for revision surgery are primarily driven by the specific failure mechanisms inherent to each material. Titanium mesh demonstrates high biological stability but is significantly limited by mechanical complications, specifically implant exposure and skin erosion. Studies by Thien (2015) show that titanium often requires removal due to these wound-related issues, with exposure rates reaching up to 13.9–17.0%, whereas PEEK implants show much higher survival rates in these categories, often as low as 1.3–4.2%. While PEEK is more prone to subgaleal fluid collection, this rarely leads to implant removal compared to the persistent skin breakdown seen with titanium^21^.

In contrast, the revision of autologous bone and PMMA is more frequently necessitated by biological failures rather than mechanical exposure. Autologous bone faces the highest risk of failure due to bone resorption, a complication exclusive to natural grafts that often leads to structural instability. Furthermore, Matsuno (2006) reported that autologous bone had the highest infection-related removal rate at 25.9%, significantly higher than titanium’s 2.6% in the same cohort. While PMMA provides a low-cost alternative, it remains susceptible to infection-related reoperations, with some studies like Wesp (2022) reporting a 20.9% reoperation rate for PMMA compared to just 2.6% for titanium-enhanced implants. Overall, the sources suggest that while synthetic custom-made materials like PEEK and certain bioceramics (HA) offer the best long-term survival by minimizing both infection and exposure, titanium mesh remains a faster, more stable option despite its risk to the overlying scalp^23^.

### Neurological outcomes

The included studies indicate that cranioplasty is a critical intervention for neurological and functional recovery, regardless of the material used for it. In large-scale assessments, titanium mesh showed a mean neurological improvement of 0.32 on the Glasgow Outcome Scale (GOS) when comparing preoperative status to the 3-to-6-month follow-up. Direct comparison between titanium and polyetheretherketone (PEEK), clinical data consistently show functional outcomes were similar. However, specialized materials like hydroxyapatite (HA) may offer greaer potential for recovery; one randomized trial demonstrated that 43% of HA patients achieved neurological improvement, a significantly higher proportion than the 26.3% observed in the titanium group.

The restoration of brain function is typically assessed across a timeline ranging from discharge to 12 months post-implantation, focusing on the resolution of neurological deficits once the skull’s integrity is restored. Ultimately, while material choice impacts complication risks, the sources suggest that successful restoration of intracranial pressure dynamics is the primary driver of functional success, provided that the surgical intervention is not undermined by major postoperative infections or exposures.

### Cosmetic and patient satisfaction

Clinical outcomes across several studies indicate that cosmetic results and patient satisfaction remain high regardless of the reconstruction material used, with no statistically significant difference generally found between titanium and alternatives like PEEK or autologous bone. For instance Chen (2023) both utilized the Rosenthal Scale (1–5) to measure aesthetic success, with the latter reporting a p-value of 1.000, indicating total parity in satisfaction among titanium, PEEK, and composite bone cement cohorts^5^. Similarly, Saxena (2023) found that 100% of patients across titanium, PEEK, and bone flap groups rated their cosmetic outcomes as "Excellent" or "Good" on a four-tier scale^25^. While custom-made implants generally facilitate superior contour restoration, some researchers suggest that titanium may be linked to accelerated scalp atrophy over time, potentially impacting long-term aesthetic stability. Furthermore, Zegers (2017) noted a Glasgow Benefit Inventory (GBI) score of 0, suggesting that from a patient’s quality-of-life perspective, there was no perceived therapeutic difference between titanium and other patient-specific implants^33^.

### Operative Metrics

The studies indicate that operative metrics, specifically surgical duration and blood loss, are influenced by the specific fabrication and molding requirements of the chosen material. Titanium mesh is frequently is used for its operative efficiency; manually shaped titanium allows for rapid intraoperative manipulation of the mesh sheet, while preformed CAD/CAM titanium further reduces the need for intraoperative contouring. In contrast, materials like hydroxyapatite (HA) and polymethyl methacrylate (PMMA) often require extensive intraoperative preparation, such as the "freehand molding" of PMMA or the complex suture fixation required for HA, which increases surgical time. The most significant outlier in operative duration is autologous bone cranioplasty, which consistently demonstrates the longest surgical times, often exceeding three hours in up to 70% of cases. This prolonged duration is primarily due to the necessity of harvesting the graft from a donor site, such as split calvarial grafts that require surgeons to meticulously harvest, carve, and assemble bone strips to span the defect. Consequently, autologous procedures not only demand more time but are also associated with higher intraoperative blood loss (e.g., 257.6 mL for autologous vs. 200.8 mL for titanium) due to the added complexity of the donor site surgery.

### Infection and wound related complications

Our study highlights a distinct trade-off between infection resistance and mechanical wound stability, as titanium mesh generally shows lower infection rates than autologous bone or PMMA but a much higher propensity for implant exposure. For example, while Matsuno (2006) found a low infection rate of 2.6% for titanium compared to 25.9% for autologous bone^23^, Yeap (2019) and Thien (2015) observed that titanium was far more likely to suffer from skin erosion and exposure, with reported rates between 13.9% and 17.0%^21,31^. This complication is frequently linked to long-term scalp atrophy and the mechanical edges of the mesh, whereas PEEK and autologous bone significantly reduce this risk, reporting exposure rates as low as 1.3% to 5.7%. However, PEEK introduces a unique wound-related challenge; subgaleal fluid collection (SFC). Studies indicate this complication is much more prevalent in PEEK cohorts than in titanium cohorts, with a significant odds ratio of 5.589. Ultimately, the failure mechanisms differ by material: while titanium mesh is primarily removed due to late-stage mechanical exposure, autologous grafts and PMMA are more frequently revised due to biological resorption or deep-seated infection.

In this systematic review and meta-analysis, titanium cranioplasty was associated with a lower risk of postoperative infection compared with PEEK implants. This association remained consistent after sensitivity analysis excluding a study with a very small titanium cohort, supporting the robustness of the findings. Several factors may contribute to the observed difference in infection risk between titanium and PEEK implants. Titanium has well-established biocompatibility and osseointegration properties, which may facilitate improved soft tissue adherence and reduced dead space. In contrast, PEEK implants, while advantageous in terms of radiolucency and customizability, may be more susceptible to bacterial colonization due to surface characteristics and limited osseointegration. From a clinical perspective, the lower infection risk observed with titanium implants may be particularly relevant in patients with prior infection, multiple comorbidities, or compromised soft tissue coverage. Nevertheless, implant selection should remain individualized, considering defect characteristics, cosmetic priorities, and surgeon experience.

### Meta-analysis

Meta-analysis was restricted to postoperative infection because other clinically relevant outcomes, including implant exposure and reoperation, were inconsistently reported across studies, with substantial variability in definitions and follow-up duration, precluding meaningful quantitative synthesis.

### Risk of Bias Assessment

Although most included studies were assessed as low risk of bias, the overall certainty of evidence for the primary outcome is limited by the predominance of retrospective designs and insufficient reporting of blinding in several studies. Therefore, the results of the meta-analysis should be interpreted within the context of moderate-quality evidence.

### Limitation

Several important limitations of this systematic review and meta-analysis should be acknowledged. First, although this review included a relatively large number of comparative studies, quantitative synthesis was feasible only for postoperative infection outcomes in the titanium versus PEEK comparison. Other clinically relevant endpoints, including implant exposure, reoperation rates, cosmetic outcomes, and neurological recovery, were reported heterogeneously across studies, with substantial variability in definitions, assessment tools, and follow-up durations. This inconsistency precluded meaningful meta-analysis of these secondary outcomes and necessitated narrative synthesis.

Second, the overall level of evidence was limited by the predominance of observational and retrospective study designs, most of which were single-center experiences with relatively small sample sizes. Although several randomized or prospective studies were included, inadequate reporting of randomization procedures and blinding was common, introducing potential selection and performance bias. In particular, blinding of outcome assessors was infrequently reported, which may have influenced the assessment of subjective outcomes such as cosmetic satisfaction and functional recovery.

Third, substantial clinical heterogeneity existed among included studies with respect to patient populations, indications for decompressive craniectomy, timing of cranioplasty, implant fabrication techniques (hand-shaped versus patient-specific implants), perioperative antibiotic protocols, and surgeon experience. These factors are known to influence complication rates and may have contributed to the observed between-study heterogeneity in infection outcomes, even within the titanium versus PEEK comparison.

Fourth, several outcomes of interest, such as implant exposure and subgaleal fluid collection, are inherently time-dependent complications. Differences in follow-up duration and incomplete reporting of time-to-event data limited the ability to assess long-term implant survival and delayed failure mechanisms in a standardized manner.

Finally, despite efforts to perform a comprehensive literature search across multiple databases without restrictions on publication year or language, the possibility of publication bias and selective outcome reporting cannot be entirely excluded. As a result, the findings of this meta-analysis, particularly those derived from a limited number of comparative studies, should be interpreted with appropriate caution.

### Future directions

Future research in cranioplasty should prioritize well-designed, adequately powered prospective studies with standardized outcome definitions and reporting frameworks. In particular, multicenter randomized or pragmatic comparative trials directly evaluating titanium, PEEK, hydroxyapatite, and other commonly used materials are needed to clarify material-specific risks across a broader range of clinically relevant outcomes beyond infection alone.

Standardization of outcome reporting represents a critical unmet need. Uniform definitions for postoperative infection, implant exposure, reoperation, and subgaleal fluid collection, as well as consistent use of validated neurological and cosmetic assessment tools, would substantially improve comparability across studies and enable more robust quantitative synthesis. Time-to-event analyses and longer follow-up periods are especially important for accurately capturing delayed mechanical failures, such as titanium mesh exposure and autologous bone resorption.

Future studies should also explore patient- and defect-specific factors that may modify the relationship between implant material and clinical outcomes, including scalp thickness, defect size and location, prior infection, comorbidities, and soft tissue quality. Subgroup analyses based on these variables may help refine individualized implant selection strategies rather than relying on a single “best” material.

Advances in implant surface engineering, additive manufacturing, and hybrid materials may offer opportunities to combine the favorable biological properties of titanium with improved soft tissue compatibility and reduced mechanical irritation. Comparative studies evaluating newer implant designs and surface modifications are therefore warranted.

Finally, incorporation of patient-reported outcome measures and cost-effectiveness analyses into future trials would provide a more comprehensive assessment of cranioplasty success from both clinical and health system perspectives.

## Conclusion

This study demonstrates that titanium cranioplasty is associated with a significantly lower risk of postoperative infection compared with PEEK implants, a finding that remained consistent after sensitivity analysis and was not driven by a single small-sample study. However, the overall evidence indicates that cranioplasty outcomes are largely determined by material-specific failure mechanisms rather than a universal superiority of any single implant. Titanium mesh offers favorable infection resistance and biological stability but is limited by a higher risk of mechanical complications, particularly implant exposure and scalp erosion. In contrast, PEEK implants show superior wound stability and lower exposure rates but are more frequently associated with subgaleal fluid collection. Autologous bone and PMMA are predominantly affected by biological failure mechanisms, including bone resorption and infection-related revision. Across materials, neurological recovery and cosmetic satisfaction appear broadly comparable, suggesting that restoration of cranial integrity itself is the primary driver of functional improvement. Taken together, these findings support an individualized, patient-centered approach to implant selection that balances infection risk, wound characteristics, defect-specific factors, and surgical priorities. While titanium may be preferable in patients at high risk of infection, no single material can be considered optimal for all clinical scenarios, underscoring the need for high-quality prospective studies with standardized outcome reporting.

## Supplementory Files

Supp_1: search strategies

Supp_2: data extraction sheet

Supp_3: ROB sheet

Supp_4: full-text screening sheet

## Supporting information

search strategy

data extraction

rob assessmnet

full text screening

## Data Availability

data extraction of studies
risk of bias assessment
search strategies

## References

1. Yang, J. et al. Evaluation of titanium cranioplasty and polyetheretherketone cranioplasty after decompressive craniectomy for traumatic brain injury: A prospective, multicenter, non-randomized controlled trial. Medicine (Baltimore) 99, e21251 (2020).

2. Koller, M. et al. A retrospective descriptive study of cranioplasty failure rates and contributing factors in novel 3D printed calcium phosphate implants compared to traditional materials. 3D Print. Med. 6, 14 (2020).

3. Wesp, D. et al. Analysis of PMMA versus CaP titanium-enhanced implants for cranioplasty after decompressive craniectomy: a retrospective observational cohort study. Neurosurg. Rev. 45, 3647–3655 (2022).

4. Guo, J. et al. A Retrospective Study on Subgaleal Fluid Collection After Titanium Mesh and Polyetheretherketone Cranioplasty. World Neurosurg. 194, 123538 (2025).

5. Chen, K. et al. Clinical Outcomes After Cranioplasty With Titanium Mesh, Polyetheretherketone, or Composite Bone Cement: A Retrospective Study. J. Craniofac. Surg. 34, 2246–2251 (2023).

6. Pfnür, A. et al. Exploring complications following cranioplasty after decompressive hemicraniectomy: A retrospective bicenter assessment of autologous, PMMA and CAD implants. Neurosurg. Rev. 47, 72 (2024).

7. Yao, S. et al. Outcome and risk factors of complications after cranioplasty with polyetheretherketone and titanium mesh: A single-center retrospective study. Front. Neurol. 13, 926436 (2022).

8. Soto, E., Boyd, C. J., Restrepo, R., Ananthasekar, S. & Myers, R. P. 72957 Rethinking reconstructive strategies for complex cranial defects: A 10 year review of cranioplasty outcomes. J. Clin. Transl. Sci. 5, 72–72 (2021).

9. Vargo, J. D., Przylecki, W. & Andrews, B. T. Surgical Decision-Making in Microvascular Reconstruction of Composite Scalp and Skull Defects. J. Craniofac. Surg. 31, 1895–1899 (2020).

10. Ji, T. et al. Subgaleal Effusion and Brain Midline Shift After Cranioplasty: A Retrospective Study Between Polyetheretherketone Cranioplasty and Titanium Cranioplasty After Decompressive Craniectomy. Front. Surg. 9, 923987 (2022).

11. Bandyopadhyay, T., Thapliyal, G. & Dubey, A. Reconstruction of Cranial Defects in Armed Forces Personnel - Our Experience. Med. J. Armed Forces India 61, 36–40 (2005).

12. Yang, J. et al. Evaluation of titanium cranioplasty and polyetheretherketone cranioplasty after decompressive craniectomy for traumatic brain injury: A prospective, multicenter, non-randomized controlled trial. Medicine (Baltimore) 99, e21251 (2020).

13. Shukla, Y. et al. Complications of Different Types of Cranioplasty and Identification of Risk Factors Associated with Cranioplasty at a Tertiary Care Centre: A Prospective Observational Study. Indian J. Neurosurg. 13, 027–034 (2024).

14. Rosinski, C. L. et al. A Retrospective Comparative Analysis of Titanium Mesh and Custom Implants for Cranioplasty. Neurosurgery 86, E15–E22 (2020).

15. Sahoo, N., Roy, I. D., Desai, A. P. & Gupta, V. Comparative Evaluation of Autogenous Calvarial Bone Graft and Alloplastic Materials for Secondary Reconstruction of Cranial Defects. J. Craniofac. Surg. 21, 79–82 (2010).

16. Afifi, A. M. et al. Lessons Learned Reconstructing Complex Scalp Defects Using Free Flaps and a Cranioplasty in One Stage. J. Craniofac. Surg. 21, 1205–1209 (2010).

17. Lindner, D. et al. Cranioplasty using custom-made hydroxyapatite versus titanium: a randomized clinical trial. J. Neurosurg. 126, 175–183 (2017).

18. Yang, J., Wang, J., You, C., Ma, L. & Guan, J. Predictors of complications following alloplastic cranioplasty in trauma patients: A multi-center retrospective study. PLOS One 20, e0321870 (2025).

19. Werndle, M. C., Crocker, M., Zoumprouli, A. & Papadopoulos, M. C. Modified acrylic cranioplasty for large cranial defects. Clin. Neurol. Neurosurg. 114, 962–964 (2012).

20. Clynch, A. L. et al. Cranial meningioma with bone involvement: surgical strategies and clinical considerations. Acta Neurochir. (Wien) 165, 1355–1363 (2023).

21. Thien, A., King, N. K. K., Ang, B. T., Wang, E. & Ng, I. Comparison of Polyetheretherketone and Titanium Cranioplasty after Decompressive Craniectomy. World Neurosurg. 83, 176–180 (2015).

22. Ng, Z. Y., Ang, W. J. J. & Nawaz, I. Computer-Designed Polyetheretherketone Implants Versus Titanium Mesh (TAcrylic Cement) in Alloplastic Cranioplasty: A Retrospective Single-Surgeon, Single-Center Study. J. Craniofac. Surg. 25, (2014).

23. Matsuno, A. et al. Analyses of the factors influencing bone graft infection after delayed cranioplasty. Acta Neurochir. (Wien) 148, 535–540 (2006).

24. Al-Tamimi, Y. Z. et al. Comparison of acrylic and titanium cranioplasty . Br. J. Neurosurg. 26, 510–513 (2012).

25. Saxena, V., Sahoo, N. K., Rangarajan, H. & Sehgal, A. “Bridging the Breach”: Cranioplasties Using Different Reconstruction Materials—An Institutional Experience. J. Maxillofac. Oral Surg. 22, 37–43 (2023).

26. Kohan, E. et al. Customized Bilaminar Resorbable Mesh With BMP-2 Promotes Cranial Bone Defect Healing. Ann. Plast. Surg. 74, 603–608 (2015).

27. Jeyaraj, C. P. Reconstruction of Large Calvarial Defects Using Titanium Mesh Versus Autologous Split Thickness Calvarial Bone Grafts: A Comprehensive Comparative Evaluation of the Two Major Cranioplasty Techniques. J. Maxillofac. Oral Surg. 17, 308–323 (2018).

28. Lönnemark, O., Ryttlefors, M. & Sundblom, J. Cranioplasty in Brain Tumor Surgery: A Single-Center Retrospective Study Investigating Cranioplasty Failure and Tumor Recurrence. World Neurosurg. 170, e313–e323 (2023).

29. Ji, T. et al. Subgaleal Effusion and Brain Midline Shift After Cranioplasty: A Retrospective Study Between Polyetheretherketone Cranioplasty and Titanium Cranioplasty After Decompressive Craniectomy. Front. Surg. 9, 923987 (2022).

30. Höhne, J. et al. Outcomes of Cranioplasty with Preformed Titanium versus Freehand Molded Polymethylmethacrylate Implants. J. Neurol. Surg. Part Cent. Eur. Neurosurg. 79, 200–205 (2018).

31. Yeap, M.-C. et al. Long-Term Complications of Cranioplasty Using Stored Autologous Bone Graft, Three-Dimensional Polymethyl Methacrylate, or Titanium Mesh After Decompressive Craniectomy: A Single-Center Experience After 596 Procedures. World Neurosurg. 128, e841–e850 (2019).

32. Binhammer, A., Jakubowski, J., Antonyshyn, O. & Binhammer, P. Comparative Cost-Effectiveness of Cranioplasty Implants. Plast. Surg. 28, 29–39 (2020).

33. Zegers, T., Ter Laak-Poort, M., Koper, D., Lethaus, B. & Kessler, P. The therapeutic effect of patient-specific implants in cranioplasty. J. Cranio-Maxillofac. Surg. 45, 82–86 (2017).

34. Okishev, D. N. et al. Features of modeling a polymer implant for closing a defect after decompressive craniotomy. Vopr. Neirokhirurgii Im. NN Burdenko 86, 17 (2022).

35. Kwiecien, G. J. et al. Long-term Effect of Cranioplasty on Overlying Scalp Atrophy. Plast. Reconstr. Surg. - Glob. Open 8, e3031 (2020).

36. Wesp, D. M. A. et al. A comparative observational analysis of PMMA versus CaP titanium-enhanced implants for cranioplasty after decompressive craniectomy. Brain Spine 2, 101635 (2022).

