## Supplementary material for "Titanium Mesh Versus Polyetheretherketone (PEEK) in Cranioplasty: A Systematic Review and Meta-Analysis of Complications and Clinical Outcomes": search strategy

Cranioplasty+Titanium vs PEEK(synthetic proteses)

Search strategy :

**Pubmed : 193**

(

cranioplasty[Title/Abstract]

OR "cranial reconstruction"[Title/Abstract]

OR "cranial repair"[Title/Abstract]

OR "skull reconstruction"[Title/Abstract]

OR "skull repair"[Title/Abstract]

OR "skull defect"[Title/Abstract]

OR cranioplast*[Title/Abstract]

OR "Cranial Defects"[Mesh]

)

AND

(

titanium[Title/Abstract]

OR "titanium mesh"[Title/Abstract]

OR "patient-specific titanium"[Title/Abstract]

OR "custom titanium"[Title/Abstract]

OR "Titanium"[Mesh]

)

AND

(

PEEK[Title/Abstract]

OR "polyether ether ketone"[Title/Abstract]

OR "poly-ether-ether-ketone"[Title/Abstract]

OR alloplastic[Title/Abstract]

OR synthetic[Title/Abstract]

OR PMMA[Title/Abstract]

OR "polymethyl methacrylate"[Title/Abstract]

OR "polymethylmethacrylate"[Title/Abstract]

OR "Acrylic Resins"[Mesh]

OR "porous polyethylene"[Title/Abstract]

OR Medpor[Title/Abstract]

)

**Scopus:293**

( TITLE-ABS-KEY ( cranioplasty ) OR TITLE-ABS-KEY ( "cranial reconstruction" ) OR TITLE-ABS-KEY ( "cranial repair" ) OR TITLE-ABS-KEY ( "skull reconstruction" ) OR TITLE-ABS-KEY ( "skull repair" ) OR TITLE-ABS-KEY ( "skull defect" ) OR TITLE-ABS-KEY ( cranioplast* ) ) AND ( TITLE-ABS-KEY ( titanium ) OR TITLE-ABS-KEY ( "titanium mesh" ) OR TITLE-ABS-KEY ( "patient-specific titanium" ) OR TITLE-ABS-KEY ( "custom titanium" ) ) AND ( TITLE-ABS-KEY ( PEEK ) OR TITLE-ABS-KEY ( "polyether ether ketone" ) OR TITLE-ABS-KEY ( "poly-ether-ether-ketone" ) OR TITLE-ABS-KEY ( alloplastic ) OR TITLE-ABS-KEY ( synthetic ) OR TITLE-ABS-KEY ( PMMA ) OR TITLE-ABS-KEY ( "polymethyl methacrylate" ) OR TITLE-ABS-KEY ( polymethylmethacrylate ) OR TITLE-ABS-KEY ( "porous polyethylene" ) OR TITLE-ABS-KEY ( Medpor ) )

**Embase:263**

(cranioplasty:ti,ab OR 'cranial reconstruction':ti,ab OR 'cranial repair':ti,ab OR 'skull reconstruction':ti,ab OR 'skull repair':ti,ab OR 'skull defect':ti,ab OR cranioplast*:ti,ab OR 'skull defect'/exp) AND (titanium:ti,ab OR 'titanium mesh':ti,ab OR 'patient-specific titanium':ti,ab OR 'custom titanium':ti,ab OR 'titanium'/exp) AND (peek:ti,ab OR 'polyether ether ketone':ti,ab OR 'poly-ether-ether-ketone':ti,ab OR alloplastic:ti,ab OR synthetic:ti,ab OR pmma:ti,ab OR 'polymethyl methacrylate':ti,ab OR polymethylmethacrylate:ti,ab OR 'porous polyethylene':ti,ab OR medpor:ti,ab OR 'acrylic resin'/exp)

**Web of science:272**

TS=(

cranioplasty

OR "cranial reconstruction"

OR "cranial repair"

OR "skull reconstruction"

OR "skull repair"

OR "skull defect"

OR cranioplast*

)

AND

TS=(

titanium

OR "titanium mesh"

OR "patient-specific titanium"

OR "custom titanium"

)

AND

TS=(

PEEK

OR "polyether ether ketone"

OR "poly-ether-ether-ketone"

OR alloplastic

OR synthetic

OR PMMA

OR "polymethyl methacrylate"

OR polymethylmethacrylate

OR "porous polyethylene"

OR Medpor

)

**Cochrane : 5**

(cranioplasty OR "cranial reconstruction" OR "cranial repair" OR "skull reconstruction" OR "skull repair" OR "skull defect" OR cranioplast*):ti,ab,kw

AND

(titanium OR "titanium mesh" OR "patient-specific titanium" OR "custom titanium"):ti,ab,kw

AND

(PEEK OR "polyether ether ketone" OR "poly-ether-ether-ketone" OR alloplastic OR synthetic OR PMMA OR "polymethyl methacrylate" OR polymethylmethacrylate OR "porous polyethylene" OR Medpor):ti,ab,kw
